# Immune signature of patients with cardiovascular disease – in-depth immunophenotyping predicts increased risk for a severe course of COVID-19

**DOI:** 10.1101/2023.04.24.23288921

**Authors:** Manina Günter, Karin Anne Lydia Mueller, Mathew Salazar, Sarah Gekeler, Carolin Prang, Tobias Harm, Meinrad Paul Gawaz, Stella E. Autenrieth

## Abstract

**Objective:** SARS-CoV-2 infection can lead to life-threatening clinical manifestations. Patients with cardiovascular disease (CVD) are at higher risk for severe courses of COVID-19. However, strategies to predict the course of SARS-CoV-2 infection in CVD patients at hospital admission are still missing. Here, we investigated whether the severity of SARS-CoV-2 infection can be predicted by analyzing the immunophenotype in the blood of CVD patients.

**Approach and Results:** We prospectively analyzed the peripheral blood of 94 participants, including CVD patients with acute SARS-CoV-2 infection, uninfected CVD patients, and healthy donors using a 36-color spectral flow cytometry panel. Clinical assessment included blood sampling, echocardiography, and electrocardiography. Patients were classified by their ISARIC WHO 4C-Mortality-Score on the day of admission into three subgroups of an expected mild, moderate, or severe course of COVID-19. Unsupervised data analysis revealed 40 clusters corresponding to major circulating immune cell populations. This revealed little differences between healthy donors and CVD patients, whereas the distribution of the cell populations changed dramatically in SARS-CoV-2-infected CVD patients. The latter had more mature NK cells, activated monocyte subsets, central memory CD4^+^ T cells, and plasmablasts than uninfected CVD patients. In contrast, fewer dendritic cells, CD16^+^ monocytes, innate lymphoid cells, and CD8^+^ T cell subsets were detected in SARS-CoV-2-infected CVD patients. We identified an immune signature characterized by low frequencies of MAIT and intermediate effector CD8^+^ T cells in combination with a high frequency of NKT cells that is predictive for CVD patients with a severe course of SARS-CoV-2 infection on hospital admission.

**Conclusion:** Acute SARS-CoV-2 infected CVD patients revealed marked changes in abundance and phenotype of several immune cell populations associated with COVID-19 severity. Our data indicate that intensified immunophenotype analyses can help identify patients at risk of severe COVID-19 at hospital admission, improving clinical outcomes through specific treatment.

**Highlights:**

- Patients with cardiovascular disease are at higher risk of severe courses of COVID-19 and may exhibit an altered immune response
- Unsupervised data analysis revealed that patients with cardiovascular disease and SARS-CoV-2 infection showed significant changes in the abundance and the phenotype of various immune cell populations
- We identified a disease-related immune signature in patients with cardiovascular disease and SARS-CoV-2 infection associated with the severity of COVID-19
- Intensified immunophenotyping helps to identify cardiovascular patients at risk of a severe course of COVID-19 already at the early stages of the disease and might thereby improve clinical outcomes and specific treatment of COVID-19

## 1. Introduction

Coronavirus disease 2019 (COVID-19) refers to a broad spectrum of clinical manifestations caused by infection with the severe acute respiratory syndrome coronavirus type 2 (SARS-CoV-2). The severity of the disease is related to risk factors such as age, sex, and pre-existing comorbidities^1, 2^ that correlate with the immune response during acute infection ^1, 2^. SARS-CoV-2 infection ranges from asymptomatic to fatal courses. In patients recovering from a non-severe infection, the immune system responded to SARS-CoV-2 infection with robust, broad-based, and transient regulatory features ^3–6^. In contrast, severe COVID-19 is characterized by hyperactivation of innate but also adaptive immune cells, an exuberant cytokine response, and high titers of SARS-CoV-2-specific antibodies ^4, 5, 7, 8^.

Common complications in hospitalized patients infected with SARS-CoV-2 include pneumonia, sepsis, acute respiratory distress syndrome (ARDS), and respiratory failure ^9–11^. The pathophysiology of SARS-CoV-2 is characterized by an early production of pro-inflammatory cytokines (e.g., tumor necrosis factor (TNF), IL-6, and IL-1β), often resulting in hyperinflammation ^1^^2^. This so-called cytokine storm increases the risk of vascular hyperpermeability and, if persistent, multi-organ failure and eventual death ^1^^1^.

Patients with cardiovascular disease (CVD) are critically susceptible to more severe courses of SARS-CoV-2 infection, which may lead to life-threatening complications like cardiac and pulmonary damage as the most common complications ^13, 14^. For example, an enhanced pro-inflammatory and pro-thrombotic immune response, characteristic of SARS-CoV-2 infected patients with pre-existing CVD, can trigger myocarditis or acute coronary syndrome with subsequent congestive heart failure ^15–19^. Furthermore, CVD patients are prone to ARDS and progressive respiratory failure and along with an increased risk of a pulmonary embolism due to infection-associated coagulopathy, which may cause not only acute right heart failure but also disseminated intravascular coagulation ^19^. These clinical scenarios explain the increased rate of unfavorable clinical outcomes like organ failure, admission to the intensive care unit (ICU) with rapidly progressive respiratory failure, and mortality in CVD patients with SARS-CoV-2 infection ^14, 15, 17^

CVD is characterized by alterations in inflammatory mediators like C-reactive protein (CRP), pro-inflammatory cytokines and chemokines, platelet and monocyte activation, and enhanced expression of adhesion molecules on endothelial and immune cells are critical players of atherogenesis and -progression ^20–22^. Up-regulation of the involved cytokines and chemokines and platelet activation recruits inflammatory cells like monocytes, macrophages, and dendritic cells to the arterial wall causing atherosclerotic lesions ^21^. In addition, pro-inflammatory adaptive immune cells like Th1 and Th17 cells are also critically involved in CVD, especially myocardial infarction, myocarditis, and heart failure ^23–25^. Due to a chronically enhanced pro-inflammatory immune response, patients with CVD are at risk for a severe course of COVID-19. However, strategies to identify these high-risk patients with CVD early in COVID-19 are still lacking. Thus, there is still an urgent clinical need to understand the complexity of the innate and adaptive immune system dysfunctions that underlie severe or even fatal COVID-19 in this subgroup of CVD patients to provide the best possible care upon hospitalization and improve clinical outcomes. In the present study, we aimed to identify the critical immune system components to predict a severe course of COVID-19 in CVD patients. Accordingly, we performed in-depth analyses of innate and adaptive immune cells and cytokines in the peripheral blood of CVD patients with and without acute SARS-CoV-2 infection. Our study revealed that a complex but specific immune signature is associated with the severity of COVID-19 in patients with CVD and can predict the course of the disease already upon first admission to the hospital.

## 2. Methods

### Study design, participants, and assessment of clinical parameters

From March to April 2020, we prospectively studied a consecutive cohort of 94 participants at the Department of Cardiology and Angiology of the University Hospital Tübingen, Germany. Of these, 37 consecutive patients with pre-existing CVD and symptomatic, acute SARS-CoV-2 infection (CVD+SARS-CoV-2) were admitted to our emergency department. Twenty patients with pre-existing stable CVD without any infections were matched for the group of CVD+SARS-CoV-2 patients. 37 healthy donors (HD) served as controls (Table 1 and Tables S1-2). All patients underwent clinical and cardiac assessment, including echocardiography, electrocardiography, concomitant medication, comorbidities, and blood sampling for routine laboratory parameters within 12 hours of admission. SARS-CoV-2 infection was diagnosed by RNA detection from nasopharyngeal secretions with real-time reverse transcriptase polymerase chain reaction. Pre-existing CVD was defined as stable coronary artery disease, which had been determined by coronary angiography and diagnosed when there was luminal stenosis of one or more coronary vessels >25-50% diameter before hospital admission. Respiratory failure was defined by a Horovitz Index (HI) ≤ 200 mmHg. ^23^

The inclusion criteria for our study were confirmed CVD with or without SARS-CoV-2 infection and an age ≥18 years. Exclusion criteria were other viral or bacterial infections and malignancies. The study was approved by the local ethics committee (240/2018BO2) and complied with the Declaration of Helsinki and the Good Clinical Practice Guidelines on the approximation of the laws, regulations, and administrative provisions of the Member States relating to the implementation of good clinical practice in the conduct of clinical trials on medicinal products for human use. Written informed consent was obtained from each patient.

N terminal-pro-B-type natriuretic peptide (NT-pro-BNP, >300 ng/L), high sensitive troponin I (hs TNI, >37 ng/L), and C-reactive protein (CRP, >0.5 mg/dL) were classified as elevated laboratory markers of myocardial and inflammatory distress. Echocardiographic parameters included left and right ventricular function, right ventricular dilatation, presence of tricuspid valve regurgitation, and pericardial effusion according to current guidelines.^24, 25^

### Isolation of peripheral blood mononuclear cells

Blood samples of patients with CVD (including SARS-CoV-2 infection) and HD were collected in CPDA monovettes and were processed within 4 hours after blood collection. Peripheral blood mononuclear cells (PBMCs) were isolated using SepMate tubes (Stem Cell Technologies) according to the manufacturer’s instructions by density gradient centrifugation at 1200 x g for 10 min at room temperature with 25 mL cell suspension (blood diluted with PBS 1:1) stacked on 15 mL Biocoll separation solution (Biochrom). The clear supernatant containing the PBMCs was decanted and washed three times with PBS+2% FCS. PBMCs were frozen as aliquots of 1-2×10^7^ cells in RPMI1640 containing 20% FBS and 10% DMSO at −150°C until further use.

### Flow Cytometry staining

PBMCs were thawed in a water bath at 37°C, followed by adding 10 ml RPMI (Sigma) containing 50 KU DNaseI (Merck). Cells were centrifuged at 400 x g for 5 minutes at room temperature. The supernatant was discarded, and the cells were resuspended in 5 ml RPMI containing 200 KU DNaseI and incubated for 20 minutes at 37°C. The cell numbers were determined by trypan blue exclusion using a Neubauer counting chamber. Cells were washed once with PBS, and 3×10^6^ cells were stained with LIVE/DEAD Fixable Blue Dead Cell Stain Kit (Thermo Fisher Scientific) according to the manufacturer’s instructions to exclude dead cells (final dilution 1:1000). All following washing and incubation steps were performed with Cell Staining Buffer (BioLegend). Blocked Cells were incubated with Human True StainFcX (BioLegend) at room temperature for 10 minutes to prevent unspecific binding of antibodies to Fc receptors. After that, extracellular staining was performed for 1 hour at 4°C using the antibodies listed in Table S3, in a final volume of 100 µl containing 5 µl True-Stain Monocyte Blocker (BioLegend) to block non-specific binding of PE/Dazzle594, PE/Cy5, PE/Cy7, APC/Cy7, and APC/Fire750 to monocytes. Cells were washed twice with staining buffer, and at least 1.5×10^6^ cells were acquired using an Aurora (Cytek Bioscience) with the SpectroFlow software. Unmixing was performed in SpectroFlow using cell- or bead-based single-stain controls and unstained cells for autofluorescence subtraction.

### Flow Cytometry data analysis

We performed a classical gating strategy (Supplemental Figure 1) for quality control of the 36-color panel and unsupervised data analysis described below using OMIQ software (Dotmatics, Boston, MA, USA). First, the compensation and the scaling were set, followed by a subsampling to 1.5×10^6^ events/sample. FlowAI was run (flow rate: second fraction 0.1; alpha 0.01; and dynamic range: both limits with negative value removal limit 1) including time, all fluorescent channels, and autofluorescence followed by gating and subsampling on 1×10^6^ flowAI passed cells/group (HD, CVD or CVD+SARS-CoV2). The data were normalized using fdaNorm for all fluorescent channels, followed by adjusting the scaling where needed and gating on and subsampling of living 5×10^5^ CD45^+^ cells per group. Subsequently, dimensionality reduction analysis was performed using Uniform Manifold Approximation and Projection (UMAP) to visualize the different sub-populations of the cells.^25^ UMAP settings were as follows: all files used, all fluorescent parameters were used except Live/Dead, CD45 and autofluorescence, Neighbors = 80, Minimum Distance = 0.7, Components = 2, Metric = Euclidean, Learning Rate = 1, Epochs = 200, Random Seed = 5733, Embedding Initialization = spectral. Following the UMAP^26, 27^ analysis, FlowSOM^27^ was run to cluster the data. FlowSOM settings were as follows: all files used, clustering features CD16, CD11c, CD56, CD8, CCR7, CD123, IgD, CD3, CD20, IgM, IgG, CD28, CD141, CD57, CD14, TCRγδ, CD25, CD4, CD27, CD1c, CD19, CD127, HLA-DR, CD38, umap_1, umap_2, xdim = 25, ydim = 25, # of training iterations = 10, Distance Metric = euclidian, consensus metaclustering with k = 100, Random Seed = 3919. A heatmap was generated with the metaclusters obtained from FlowSOM and clustered hierarchically using the samples from HD with the medians of all surface markers except for LIVE/DEAD, CD45, and autofluorescence with a euclidean row distance and a ward row leakage to assign the metaclusters to the cell populations.

### Determination of plasma levels of cytokines/chemokines (LEGENDPlex)

Two LEGENDPlex (Inflammation Panel 1 and Pro-inflammatory Chemokine Panel; BioLegend, San Diego, California, USA) were performed to quantify the concentrations of several chemokines and cytokines in human plasma. Sixty-nine frozen plasma samples were analyzed, consisting of 27 HD, 15 CVD, and 27 SARS-CoV-2-infected CVD patients. The assays were performed according to the manufacturer’s manual. A FACS Lyric (BD Biosciences, Franklin Lakes, New Jersey, USA) was used for the measurement. Data analysis was performed with the LEGENDPlex Data Analysis Software (BioLegend, San Diego, California, USA).

### Statistical analysis

We determined clinical and laboratory baseline characteristics in relation to measured immune cell phenotypes, marker expression, and clinical outcome. Continuous, not normally distributed variables are expressed as median with standard deviation and were compared using unpaired two-tailed Mann–Whitney *U* test for two-group comparison or Kruskal Wallis non-parametric test with Dunn’s post-test for three-group comparison. Categorical data are presented as total numbers and proportions and were analyzed by chi-squared test. Correlation analysis was performed by Spearman rank correlation coefficient r. To summarize, correlations of essential parameters in CVD patients with and without SARS-CoV-2 infection matrices were generated using Rstudio “Corrplot,” displaying correlations of cytokines with or without cell populations from the unsupervised data analysis. Spearman’s ρ is colored, and color intensity and size are plotted proportionally to correlation coefficients.

The “CombiROC” package was used in Rstudio to determine the optimal combination of cell populations to differentiate mild from severe or moderately SARS-CoV-2-infected CVD patients.^28, 29^ Orthogonal partial least-square discriminant analysis (OPLS-DA) was performed using the “MetaboAnalyst” package in RStudio.

Comparisons were considered statistically significant if the two-sided p-value was <0.05. Statistical analysis was performed using Prism Software Version 9.4.1 (GraphPad).

## Results

### Clinical characteristics of patients with cardiovascular disease and symptomatic acute SARS-CoV-2 infection

We prospectively studied a cohort of 94 participants from February to April 2020. This cohort consisted of 20 patients with pre-existing CVD and 37 with pre-existing CVD and symptomatic acute SARS-CoV-2 infection. 37 HD served as controls. The baseline characteristics and demographics of the overall cohort are given in **Table 1**. The population’s median age was 58 (IQR 42–74) years, and 45 (47.9%) patients were men. Detailed information on every subgroup of the cohort is given in Table S1. Out of 37 CVD+SARS-CoV-2 patients, 20 showed respiratory failure with HI ≤ 200 mmHg, while 11 (29.7%) were admitted to the ICU due to progressive respiratory, circulatory, or multi-organ failure. CVD+SARS-CoV-2 patients were further stratified by their ISARIC-WHO-4C-Mortality-Score into a group of expected mild, moderate, or severe course of COVID-19, patients’ characteristics stratified by ISARIC-WHO-4C-Mortality-Score are depicted in Table S2. Patients with expected mild COVID-19 were significantly younger (p=0.016) than those with an expected moderate or severe disease course. Interestingly, cardiovascular risk factors were equally distributed among the three groups, whereas laboratory parameters like lymphocyte count, CRP, and interleukin-(IL-)6 levels showed significant alterations.

### Characterization of immune cell subsets in peripheral blood using a 36-color spectral flow cytometry panel

Frozen peripheral blood mononuclear cells (PBMCs) from the described patient cohort were stained with a 36-color antibody panel **(Suppl. Figure 1**) related to a previously published panel ^30^ and measured with a spectral flow cytometer (Aurora, Cytek) to characterize all common immune cell subsets (**Fig. 1A**). Data analysis was performed classically by manual gating (**Suppl. Figure 2**) as well as by unsupervised analysis using Uniform Manifold Approximation and Projection (UMAP)^26^ for dimensionality reduction followed by clustering using self-organized map (FlowSOM)^27^ (**Figure 1C**). Already in the UMAP visualization differences were seen between the concatenated files of the different groups of the cohort (**Figure 1B**). FlowSOM clustering was performed with 100 metaclusters (MCs) to identify cell populations with lower abundance, such as cDC1. 13 of these 100 MCs were excluded for further analysis as the percentage of cells in these MCs was less than 0.01. FlowSOM clustering in combination with UMAP plots displaying the expression of one lineage marker on the color-coded x-axis (**Suppl. Figure 3**) and a clustered heatmap showing the median expression of the analyzed markers (**Figure 1D**) was used to assign the obtained 87 clusters to distinct cell populations (**Figure 1C**). Finally, the 36-color staining followed by unsupervised data analysis revealed the discrimination of 40 manually assigned specific cell populations reflecting the manual gating strategy. These cell populations include B cell (lilac), CD4^+^ (orange) and CD8^+^ T cell (blue), NK cell (green), monocyte (pink), and dendritic cell subsets (cDCs, violet), innate lymphoid cells (ILCs, light green) and MAIT cells (turquoise) as well as basophils (yellow) and also neutrophils (purple).

**Figure 1:**
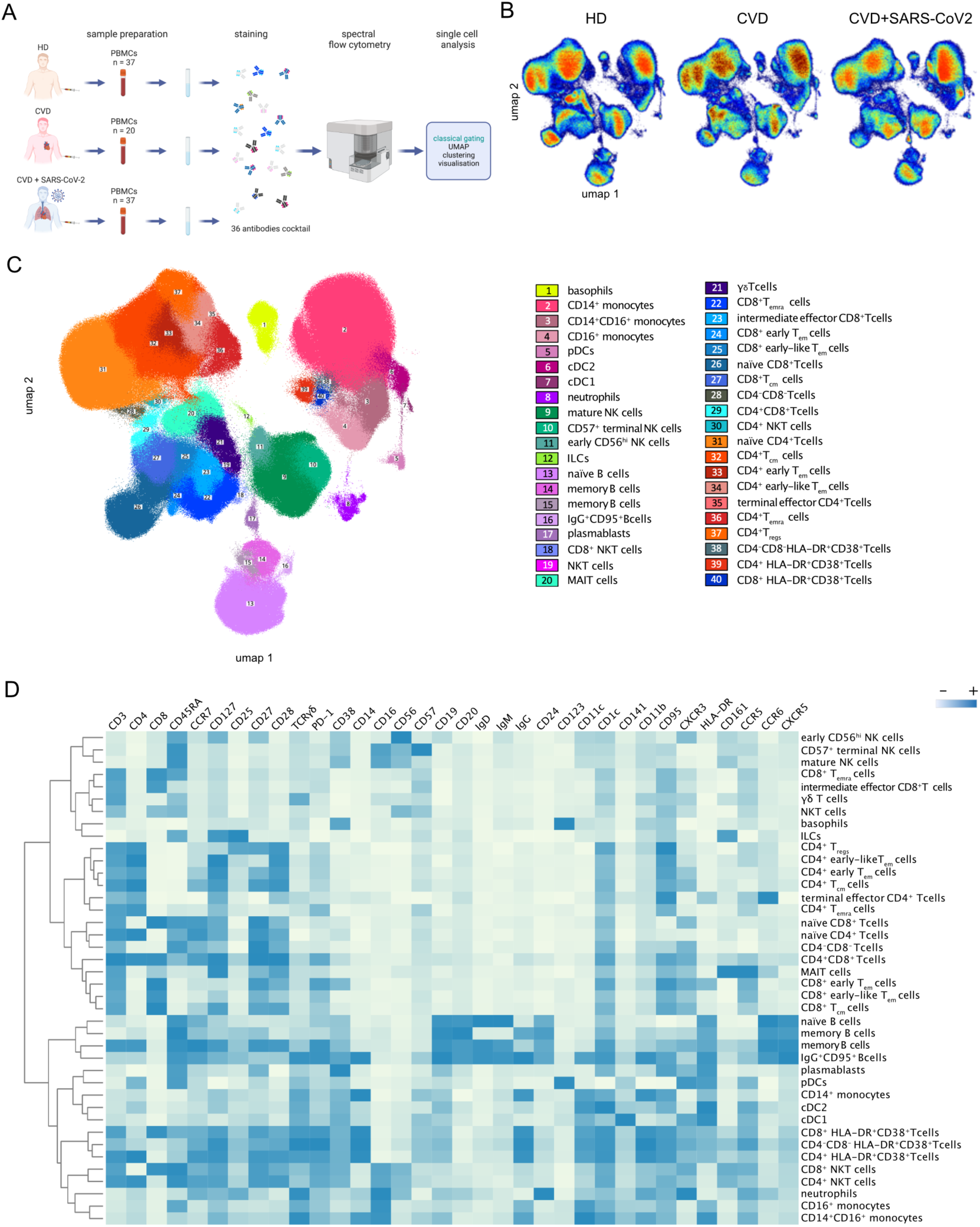
UMAP dimensional data analysis of PMBCs using a 36-color antibody panel. **A** Experimental design **B** High-dimensional data analysis of PBMCs from healthy donors (HD, 37), patients with cardiovascular disease (CVD, 20), and patients with CVD and SARS-CoV-2 infection (CVD + SARS-CoV2 patients, 37) displayed in two UMAP dimensions. The density plots show concatenated events from all of the indicated samples. **C** 40 FlowSOM clusters projected onto two UMAP dimensions. The overlay plot shows concatenated events from all samples. FlowSOM clusters were assigned to depicted cell populations based on their marker expression (see heatmap in **D**). **D** Hierarchically clustered heatmap displaying the marker expressions of manually labeled 40 FlowSOM clusters from 37 concatenated HD samples. The marker expression intensity is displayed on a scale from white (negative) to blue (positive). Each column’s max and min are mapped to this scale, so the values change between markers; consequently, the scale is labeled with – and + to indicate relative magnitude.

### SARS-CoV2-infected CVD patients showed significant differences in the distribution and the phenotype of immune cell populations compared to uninfected CVD patients

Next, we assessed the differences in the abundance of the 87 MCs obtained by FlowSOM (annotated to 40 different cell populations as described in **Fig. 1**) in PBMCs from HD and CVD patients with and without SARS-CoV2 infection (**Suppl. Figure 4**). Comparison of HD and uninfected CVD patients revealed a reduced frequency of clusters assigned to γδ T cells, MAIT cells, naïve CD8^+^ T cells, early-like effector memory CD4^+^ T cells (CD4^+^ early-like T_em_), CD4^-^CD8^-^ T cells, and naïve B cells in CVD patients. In contrast, clusters representing more innate inflammatory cells like CD14^+^ monocytes, neutrophils, mature NK cells, and CD8^+^ central memory T cells (CD8^+^ T_cm_) were increased in frequency in CVD patients compared to HD (**Suppl. Figure 5**).

During symptomatic, acute SARS-CoV-2 infection in hospitalized CVD patients, we observed significant changes in their immune signature compared to uninfected CVD patients, particularly a greater proportion of mature NK cells (MC33), CD14^+^CD16^+^ monocytes (MC11, MC35), activated populations of CD14^+^CD45RA^+^ monocytes (MC27, MC50, MC63), CD4^+^ T_cm_ cells (MC97) expressing CD38^+^, CD8^+^ early T_em_ cells (MC60) expressing HLA-DR^+^CD38^+^PD-1^+^ as well as plasmablasts (MC31, MC39) (**Figure 2B**). In contrast, reduced proportions of ILCs (MC62), dendritic cell subsets cDC1 (MC14) and cDC2 (MC13), CD16^+^ monocytes (MC23, MC34), less activated/mature CD14^+^ monocytes (MC28, MC45, MC59), as well as CD4^+^CD8^+^ T cells (MC76), naïve, central memory, and early effector-memory CD8^+^ T cells (MC67, MC68, MC61) and CD4^+^ T_regs_ (MC100) were observed in SARS-CoV2-infected compared to uninfected CVD patients (**Figure 2**). To complement our unsupervised analysis, we applied manual gating (**Suppl. Figure 2**) for immune cell subsets contributing to SARS-CoV-2 infection. Similar to the unsupervised data analysis results, we found increased numbers of CD14^+^CD16^+^ intermediate monocytes and plasmablasts and, although not found in the unsupervised data analysis, increased numbers of CD4^+^ T_emra_ cells in SARS-CoV2-infected CVD patients (**Suppl. Figure 6**). The numbers of ILCs, dendritic cell subsets pDCs, cDC1, and cDC2 (not statistically significant), CD16^+^ monocytes, as well as naïve, central memory, and early effector-memory CD4^+^ and CD8^+^ T cells and CD4^+^ T_regs_, were reduced in SARS-CoV-2 infected compared to uninfected CVD patients. Moreover, fewer numbers of T helper 17 (T_h17_) and follicular T helper (T_fh_) cells and γδ T cells were detected upon SARS-CoV-2 infection.

**Figure 2:**
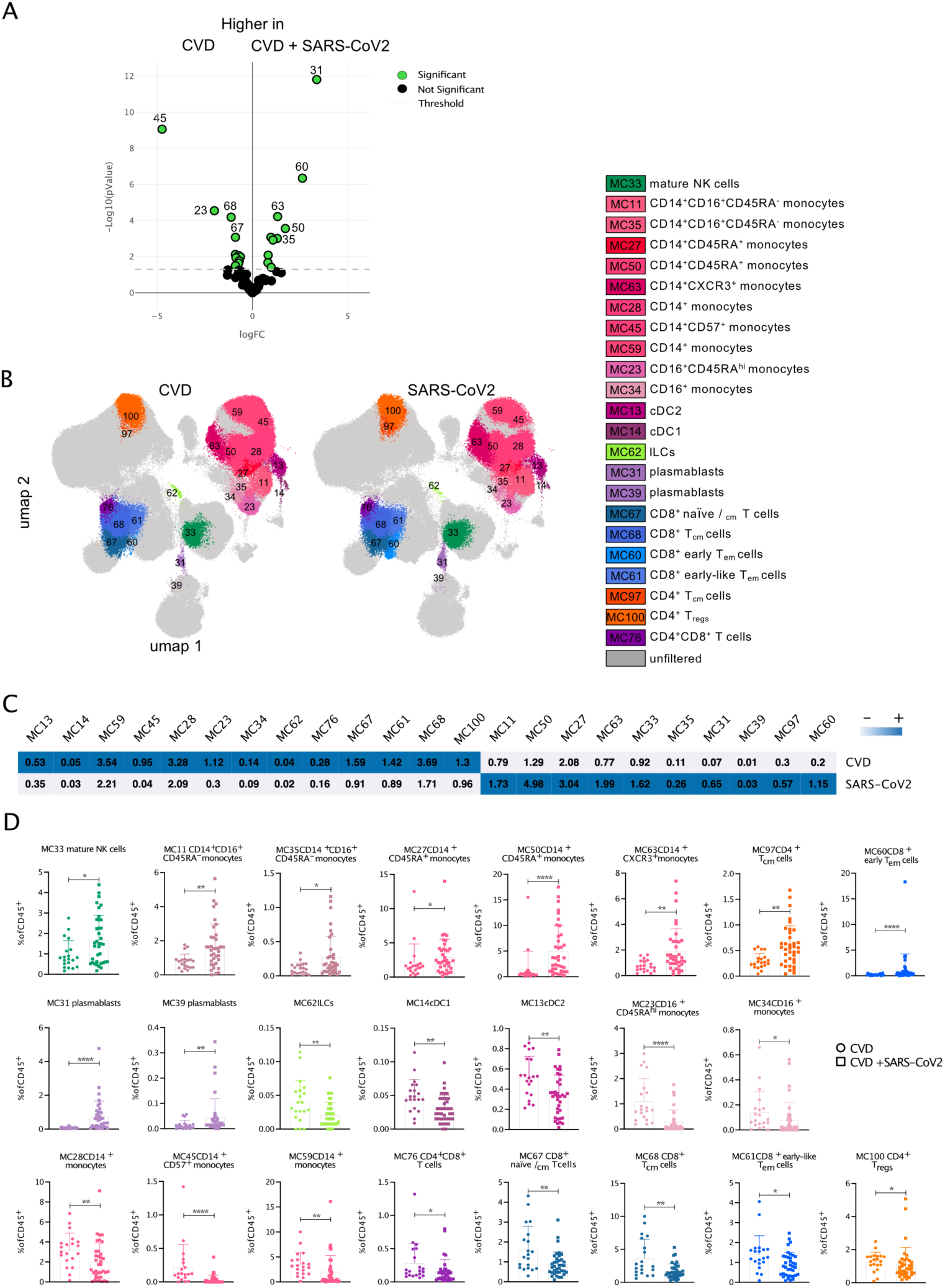
Differences between immune cell populations of CVD patients compared to SARS-CoV2-infected CVD patients. **A** Volcano plot comparing the frequency of cells in 87 FSOM clusters from CVD and SARS-CoV2-infected CVD patients using edgeR. Significant different clusters are depicted in green. **B** The overlay UMAP-plots show concatenated events from CVD (20) and SARS-CoV2-infected CVD (37) samples highlighting the significantly different clusters from A. **C** Clustered heatmap displaying the frequency of cells in the FSOM clusters from concatenated samples from CVD (upper row) and SARS-CoV2-infected CVD patients (lower row). The frequency is displayed per row on a scale from white (low) to blue (high). **D** Box plots show the abundance of the depicted MC with assigned cell populations of the individual samples from CVD and SARS-CoV2-infected CVD patients. Data were analyzed using Mann-Whitney non-parametric test. Significant differences between the two patient cohorts are marked with stars (* > 0.05; ** > 0.01; *** > 0.001; **** > 0.0001). MC, metacluster

In addition to changes in the proportion of immune cells, we also observed phenotypical changes in SARS-CoV-2 infected CVD patients compared to uninfected CVD patients (**Suppl. Figure 7**). In particular, an overall activation of innate immune cells was observed. For example, the expression of CD38, CD95, and CCR5 was significantly increased on NK cell subsets of SARS-CoV-2 infected CVD patients. Plasmacytoid DCs, important in viral infections due to their type I interferon response ^31^, also showed a significantly more activated phenotype by increased expression of CCR7, CD38, and CD95. Similar effects were observed for basophils and innate lymphocyte cells (ILCs) with increased expression of CD38, CD95, and CXCR3 or CD45RA, CD28, and CD25, respectively.

In contrast, the cDC subsets from SARS-CoV-2 infected CVD patients showed less expression of HLA-DR and CD38 but increased expression of CCR5, CCR7, CD11b, CD45RA, and IgG (cDC2s) (**Suppl. Figure 7**), indicating cellular activation and migration, but also an association with a reduced antigen presentation capacity. Regarding the monocyte clusters, we observed an increased migratory and activated phenotype in SARS-CoV-2 infected CVD patients, determined by the high expression of CCR7 in all monocyte subsets. However, monocyte subsets from these patients revealed a less activated phenotype than their uninfected CVD counterparts, shown by significantly less expression of HLA-DR, CD38, CD11b (on CD14^+^ monocytes), and CD45RA. Most strikingly, we found an enhanced expression of the thrombin receptor CD141 on monocyte clusters from SARS-CoV-2 infected patients, possibly aiming toward impaired blood coagulation ^32^.

Concerning the adaptive immune cell populations, a more inhibitory phenotype was observed in SARS-CoV2-infected CVD patients compared to uninfected (**Suppl. Figure 7**). The only exception was the plasmablasts. Plasmablasts revealed reduced expression of the apoptotic marker CD95 combined with increased CD38 expression, indicating a more functional and long-living phenotype upon SARS-CoV2 infection. Similar to DCs, a phenotype with decreased activation was observed in the B-cell subsets of SARS-CoV2-infected CVD patients. This was evidenced by lower CD38, HLA-DR, CCR6, CD45RA, and IgD expression. CD8^+^ and most CD4^+^ T cell subsets showed an overall increase in CD38 expression, indicating activation upon SARS-CoV2 infection. However, the inhibitory molecule PD-1 on the cell surface of CD4^+^ and CD8^+^ naïve and T_cm_ cells, CD4^+^ T_regs_, and HLA-DR^+^CD8^+^ activated T cells was increased, which was accompanied by less expression of the co-stimulatory molecule CD28 on CD8^+^ T cell subsets. CXCR3 expression was also reduced on several CD4^+^ and CD8^+^ T cell subsets of SARS-CoV-2 infected compared to uninfected CVD patients. The latter two observations hint towards an impaired migratory capacity and full T cell activation of CD4^+^ and CD8^+^ T cell subsets. In summary, our data show an infection-induced activation of innate immune cells in combination with reduced numbers and inhibitory phenotypes of DCs and adaptive immune cells in infected compared to uninfected CVD patients.

### Chemokine and cytokine profiling showed significant differences between SARS-CoV-2 infected and uninfected CVD patients

In addition to spectral flow cytometry, 25 common cytokines and chemokines were analyzed in plasma samples from HD and CVD patients without and with SARS-CoV-2 infection, the latter being further subdivided into groups with expected mild and severe course of COVID-19 based on the ISARIC-WHO-4C-Mortality-Score. In line with other studies, the pro-inflammatory cytokines IL-6 and IL-18 were significantly increased in the plasma of SARS-CoV-2 infected compared to uninfected CVD patients and HD (**Suppl. Figure 8A)** ^33, 34^. Furthermore, IL-6 was also elevated in CVD patients compared to HD and significantly increased in severe compared to mild SARS-CoV2-infected CVD patients (**Suppl. Figure 8A&B**). Interestingly, TNF, IL-1b, IL-12p70, IL-23, and IL-33 were lower in SARS-CoV-2 infected CVD patients than uninfected CVD patients and HD, indicating impaired T helper cell differentiation in this subgroup. In contrast, the chemokines CCL2 and the CXCR3 ligands CXCL9, CXCL10, and CXCL11 were markedly elevated during SARS-CoV-2 infection. Differences between mildly and severely affected CVD patients were observed; in particular, a significant increase of IL-6 and IL-8, whereas CCL17, responsible for the recruitment of T cells^35^, was significantly less abundant in severely infected CVD patients (**Suppl. Figure 8B**). Correlation analysis revealed that predefined cell clusters and chemokine/cytokine profiling showed significant associations among the subgroup of SARS-CoV-2 infected CVD patients and CVD patients without any infections, as depicted in **Suppl. Figure 8C**.

### An immune signature of SARS-CoV-2 infected CVD patients is associated with the severity of COVID-19

SARS-CoV-2 infected CVD patients were classified by their ISARIC WHO 4C-Mortality-Score on the day of admission into three subgroups of an expected mild (4C-Score <4), moderate (4C-Score 4-8), or severe (4C-Score ≥9) course of COVID-19. To determine an association between the immune signature and the severity of COVID-19, we analyzed the abundance and alterations of immune cell populations in the peripheral blood of CVD patients with expected mild, moderate, or severe course of COVID-19. The extent to which the immunophenotype within these three groups may differ was evaluated by spectral flow cytometry as described above.

Comparing the frequencies of cells in the above-defined 87 MC from patients with mild and severe or moderate and severe courses of COVID-19 revealed 19 significantly different MCs among the groups (**Figure 3A, Suppl. Figure 9**). These comprised reduced frequencies of neutrophils (MC8, MC16), MAIT cells (MC65, MC69)^36^, CD14^+^ monocytes expressing high levels of HLA-DR (MC12, MC28), and IgG^+^CD95^+^ B cells (MC25) in severe compared to mild SARS-CoV-2 infected CVD patients. In contrast, a greater proportion of mature NK cells (MC26, MC32), CD45RA^+^CD14^+^ monocytes (MC50), CD8^+^NKT cells (MC55), and several CD4^+^ and CD8^+^ T cell subsets, including CD4^-^CD8^-^HLA-DR^+^ T cells (MC49), naïve CD4^+^ T cells expressing CCR6 and CXCR3 (MC85), central memory (MC79), and effector subsets (MC51, MC87, MC74; **Figure 3**). Interestingly, the percentage of CD14^+^HLA-DR^+^ monocytes expressing high levels of HLA-DR (MC12, MC28), as well as that of CD8^+^ T_em_ (MC51) cells, was reduced in severe compared to moderate SARS-CoV-2 infected CVD patients (**Figure 3**). In addition to changes in the percentage of immune cell populations in the blood of SARS-CoV-2 infected CVD patients with different severity, we also found differences in the expression pattern of immune markers depending on disease severity (**Suppl. Figure 10**). For example, CD19 was significantly less expressed on MC that represent naïve and memory B cells (MC1, MC2, MC3, MC4, MC19, MC20, MC25, MC21) in SARS-CoV-2 infected CVD patients with a severe compared to mild and/or moderate course, indicating less B cell receptor signaling^37^. Similarly, the expression of the FcgRIII CD16 was reduced on neutrophils (MC7), CD14^+^ (MC12), CD14^+^CD16^+^ (MC11, MC18), CD16^+^ monocytes (MC37), mature NK cells (MC6, MC33), NKT cells (MC41) as well as on CD8^+^ T_em_ and T_emra_ cells (MC51, MC52, MC40, MC46) and CD4^+^ T_cm_ cells (MC79) from CVD patients suffering from severe COVID-19 compared to those with a mild course, indicating less antibody-dependent cellular toxicity and thereby less killing of virus-infected cells (ADCC ^38^). Accordingly, CD14^+^CD16^+^ monocytes from severe SARS-CoV2-infected CVD patients express less HLA-DR on their surface (MC38). The chemokine receptor CXCR3, which is generally upregulated on activated NK and T cells^39^, had lower expression levels on immune cells from CVD patients with a severe course of COVID-19 namely on neutrophils (MC8), mature NK cells (MC32; MC33), MAIT cells (MC65) and several clusters representing CD4^+^ naïve, T_cm_ and T_em_ cells (MC85, MC93, MC73, MC79, MC95, MC86). CD127 (IL7Rα) was significantly less expressed by ILCs (MC62) and CD4^+^CD8^+^ T cells (MC76) from CVD patients with severe SARS-CoV-2 infection as compared to those with mild infection (**Suppl. Figure 10**). In contrast, the latter showed more CCR7 expression (MC76), indicating an altered function of these cells. Furthermore, CD16^+^ monocytes (MC23), naïve CD4^+^ and CD8^+^ T cells (MC84, MC93, MC67) from CVD patients with a severe course of infection, expressed higher levels of CD95 and CD161 (MC84, MC70), combined with lower levels of CD28 expression at least on naïve CD4^+^ T cells (MC85). CD16^+^ monocytes (MC23), naïve CD4^+^ T cells (MC92), and CD4^-^CD8^-^HLA-DR^+^ T cells (MC49) expressed higher levels of CD141 (**Suppl. Figure 10**).

**Figure 3:**
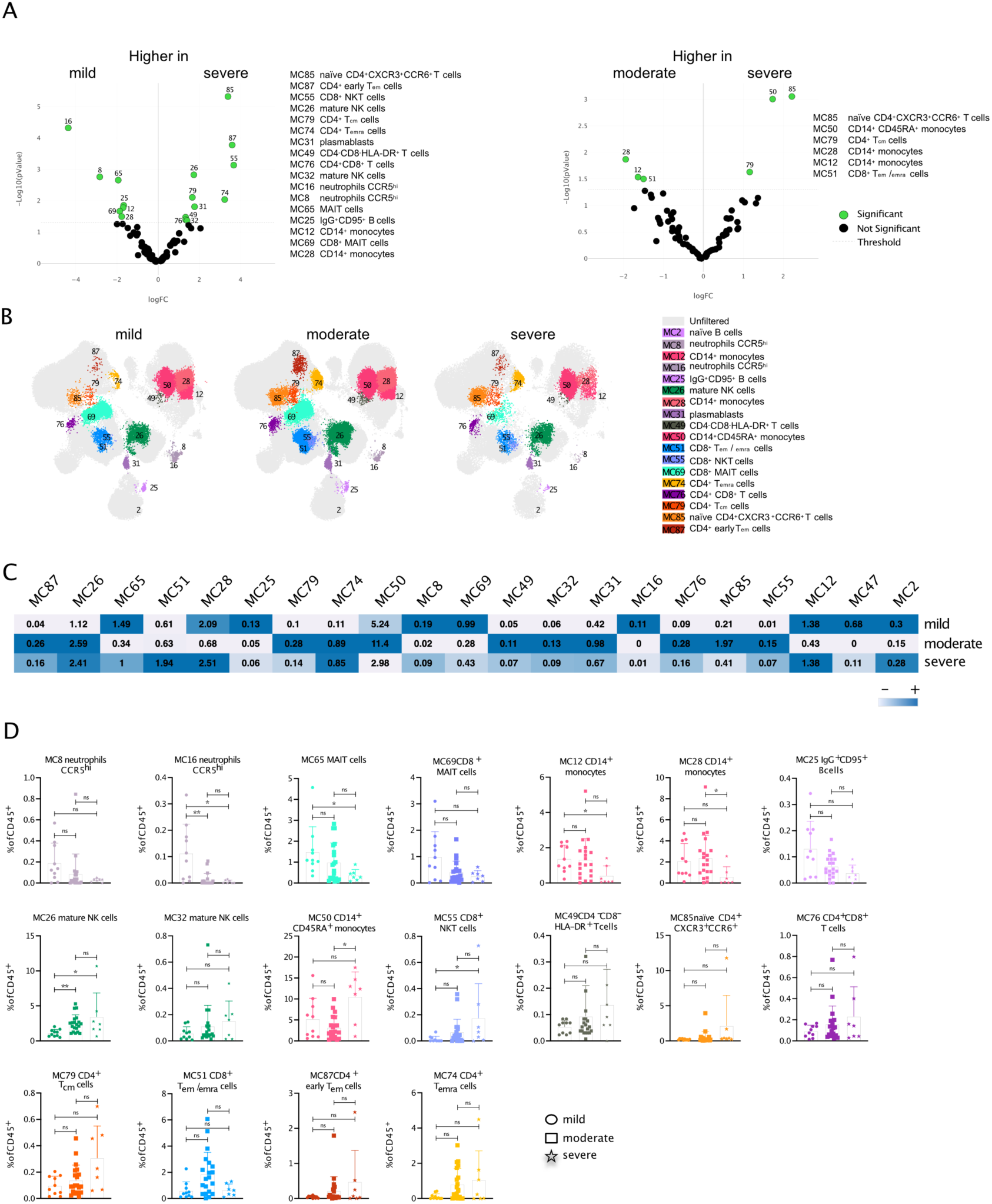
Differences between immune cell populations in mild, moderate, and severe SARS-CoV2-infected CVD patients. **A** Volcano plots comparing the frequency of cells in the 87 FSOM clusters from mild (10), moderate (20), and severe (7) SARS-CoV2-infected CVD patients using edgeR. Significant different clusters are depicted in green. **B** The overlay UMAP plot shows concatenated events from mild (10), moderate (20), and severe (7) SARS-CoV2-infected CVD patients highlighting the significantly different MCs from A. **C** Clustered heatmap displaying the frequency of cells in the FSOM clusters from concatenated samples from severe (upper row), moderate (middle row), and mild SARS-CoV2-infected CVD patients (lower row). The frequency is displayed per row on a scale from white (low) to blue (high). **D** Box plots show the abundance of the depicted MC with assigned cell populations of the individual samples. Data were analyzed using Kruskal-Wallis non-parametric test with Dunn’s post-test. Significant differences between the patient cohorts are marked with stars (* > 0.05; ** > 0.01; *** > 0.001).

In summary, severe SARS-CoV-2 infection resulted in fewer innate immune cells. Interestingly, although an increased proportion of NK cells, CD4^+,^ and CD8^+^ T cell subsets were observed in CVD patients with severe COVID-19, their expression of functional markers like CD16 and CXCR3 was impaired, indicating an altered immune response in this subgroup.

### Immune signature is predictive of severity and the course of SARS-CoV-2 infection in patients with pre-existing cardiovascular disease

Next, we looked for an immune signature that has the potential to predict the course of infection at hospital admission. We decided to use a combination of cell populations obtained through unsupervised data analysis that may provide a clear indication of disease progression through a single analysis method. To do this, we first compared the number of cells within different immune cell subpopulations in CVD patients with mild and severe courses as well as moderate and severe courses of SARS-CoV-2 infection (**Figure 4A**). Higher numbers of neutrophils, MAIT cells, and IgG^+^CD95^+^ B cells were found in mild-infected SARS-CoV-2 CVD patients. In contrast, the numbers of NKT cells, plasmablasts, CD4^-^CD8^-^HLA-DR^+^ T cells, and CD4^+^ T_emra_ cells were higher in severely infected SARS-CoV-2 CVD patients (**Figure 4A**). When comparing moderately and severely infected patients, only the intermediate effector CD8^+^ T cells, CD8^+^ NKT cells, and cDC2 were higher in moderately SARS-CoV-2-infected CVD patients (**Figure 4A&B**).

**Figure 4:**
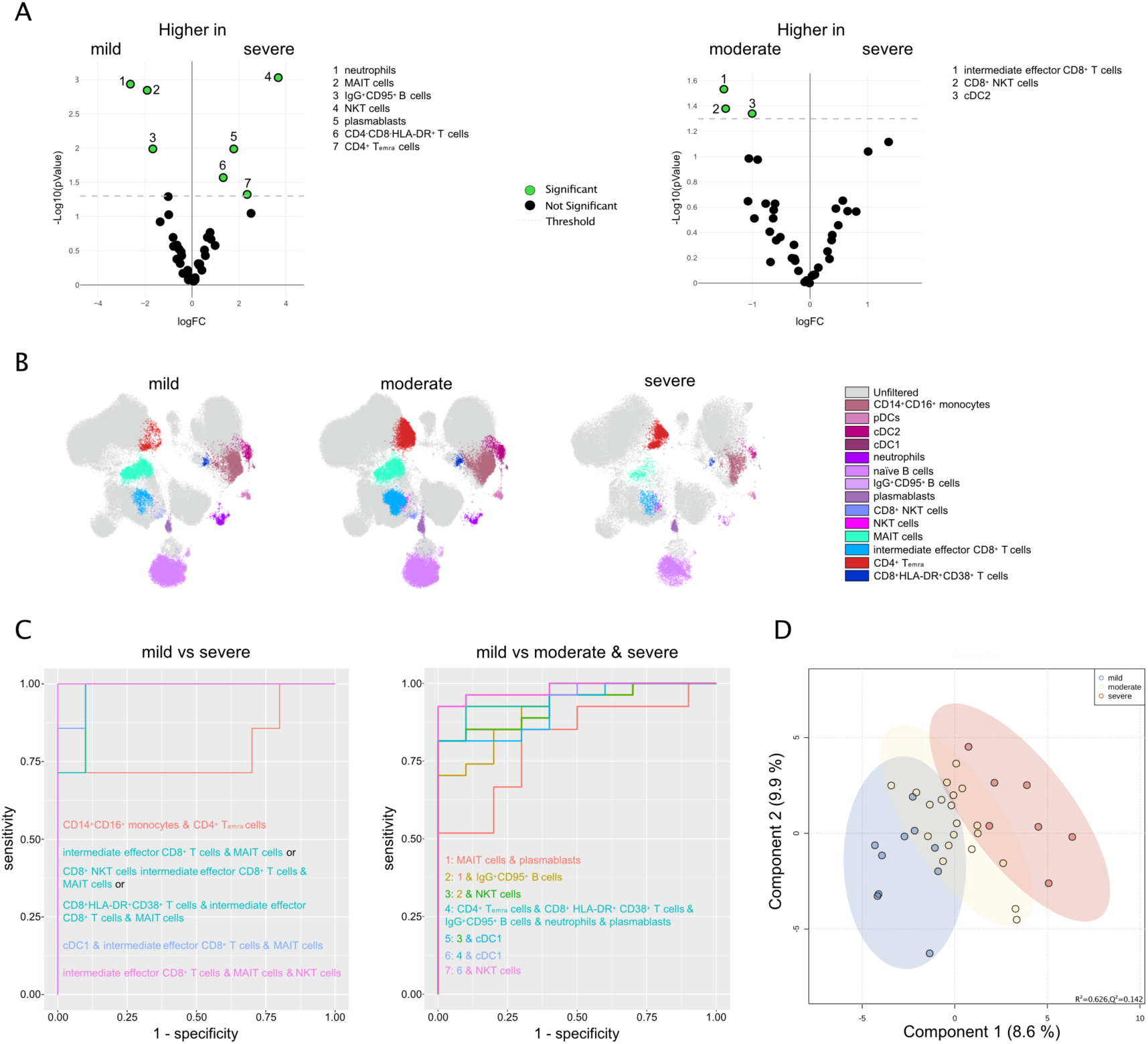
Correlation of immune cell abundancy with disease severity. **A** Volcano plot comparing the frequency of cells in the 40 assigned cell populations (from Fig. 1) from mild (10), moderate (20), and severe (7) SARS-CoV2-infected CVD patients using edgeR. Significantly different cell populations are depicted in green. **B** The overlay UMAP plot shows concatenated events from mild (10), moderate (20), or severe (7) SARS-CoV2-infected CVD patients highlighting the significantly different cell populations from A. **C** CombiROC analysis using the frequencies of the significantly different cell populations from A. Patients stratified for a mild course of infection compared either with severely stratified patients (left) or the combination of moderate and severe stratified patients (right). **D** Orthogonal partial-least square (OPLS) analysis comprising immune cell populations and cytokine/chemokine plasma levels of patients with SARS-CoV2 infection. Dots represent single study subjects and are colored by disease severity (with mild = blue, moderate = yellow, severe = red points). X-axis and Y-axis show the T score as well as the percentage of explained variance.

After screening for the most relevant differentially abundant immune cell populations in the different patient groups, we used these to search for biomarker combinations to identify subjects at high risk of developing severe COVID-19. This search was performed using the recently published CombiROC package ^20^. Since moderately infected CVD patients may also develop a potentially dramatic course of infection and long-COVID, we also looked for a combination of cell populations that could distinguish this group from the cohort with a mild course. Thus, we used three settings for combined biomarker search: comparisons of (1) mild versus severe (**Figure 4C**), (2) mild versus moderately (**Suppl. Figure 11A**), and (3) mild versus moderately and severely (**Figure 4C**) infected CVD patients. Using CombiROC for the first comparison revealed that a combination of only three cell populations, namely MAIT cells, intermediate effector CD8^+^ T cells and, NKT cells predicts a severe course of SARS-CoV-2-infected CVD patients on hospital admission with a sensitivity and specificity of 1 (pink line) (**Figure 4C** & **Table S4**). The two other comparisons, (2) and (3), did not give quite as good results. In both cases, an area under the curve (AUC) of 0.98 was achieved at best, with a specificity of 1 and a sensitivity of 0.95 and 0.9267, respectively (**Figure 4C** & **Tables S5-S6**). However, a combination of 7 cell populations was necessary in both cases. The distinction between a moderate and a mild course of infection should be possible considering CD4^+^ T, CD8^+^ and CD8^-^ NKT cells, CD8^+^HLA-DR^+^CD38^+^ T cells, IgG^+^CD95^+^ B cells, neutrophils, and plasmablasts. In the case of predicting a mild compared to a moderate or severe infection course if SARS-CoV-2 infection, a combination of CD4^+^ T_emra_, CD8^+^HLA-DR^+^CD38^+^ T cells, IgG^+^CD95^+^ B cells, neutrophils, plasmablasts, cDC1, and NKT cells is necessary.

Correlation analyses were performed between the patient groups’ most relevant differentially abundant immune cell populations (see **Figure 4A**) and the corresponding cytokine/chemokine plasma levels. Uninfected CVD patients were characterized mainly by negative correlations, e.g., cDC1s with IFN-a, IL-10, IL-12p70, TNF or CD8^+^HLA-DR^+^CD38^+^ T cells with CXCL10 (**Suppl. Figure 11B**). In contrast, SARS-CoV-2-infected CVD patients showed negative correlations of CD8^+^HLA-DR^+^CD38^+^ T cells with CCL5 or cDC2s with CXCL10, IFN-γ, and IL-10 as well as positive correlations like naïve B cells and plasmablasts with CXCL9, IL-10, and IL-18, or IFN-α with IgG^+^CD95^+^ B cells, naïve B cells, and CD8^+^ NKT cells (**Suppl. Figure 11B**). Mildly SARS-CoV-2-infected CVD patients showed positive CD8^+^ and CD8^-^ NKT cells correlations with CCL2, CXCL10, CXCL9, IFN-α, IFN-γ, and IL-10, but negative correlations of cDC1 and cDC2 with CXCL10 and IFN-α (**Suppl. Figure 11C**). Significantly fewer and also less pronounced correlations were observed in patients with a moderate course of SARS-CoV2 infection. The positive correlations of IgG^+^CD95^+^ B cells and plasmablasts with IL-10, IL-12p70, IL-18, and, in the latter case, also with IL-6 should be noted. More correlations were generally observed again in the case of a severe SARS-CoV2 infection. Here, positive correlations of naïve B cells, plasmablasts, CD8^+^ NKT cells, and neutrophils with CCL2, IFN-α, IFN-γ, IL-17, and IL-6 dominated(**Suppl. Figure 11C).** Moreover, score plots of orthogonal partial least-squares discriminant analysis (OPLS-DA) were pereformed using immune cell populations and cytokine/chemokine plasma leverls from SARS-CoV-2 infected CVD patients (**Figure 4D**). Good between group variance of patients with mild (blue) compared to severe infection (red) points out a major impact of altered immune signatures on the clinical course of the disease.

In summary, using high-resolution flow cytometry, we identified an immune cell combination consisting of MAIT cells, intermediate effector CD8^+^ T cells, and NKT cells, stratifying CVD patients at high risk for severe SARS-CoV2 infection on the day of hospital admission.

## Discussion

Our study identified a characteristic immune signature of patients with CVD and acute, symptomatic SARS-CoV-2 infection. In contrast to previous investigations, we focussed on patients with pre-existing CVD, which are at high risk of developing progressive cardiopulmonary failure due to the infection ^13–15^. To our knowledge, this is the first analysis to associate a specific immune signature of CVD patients with SARS-CoV-2 infection with the expected severity of COVID-19 during the disease. Here, we identified key immune system components critical to predicting a severe course of COVID-19 in our patient cohort through in-depth analyses of innate and adaptive immune cells ^9, 10^. Most other studies that have examined immune cell populations in the blood in SARS-CoV-2 infection have included an all-comers cohort of patients and have not stratified their analyses by concomitant comorbidities, such as diabetes, obesity, asthma, particularly the presence of CVD ^4, 5, 8, 40–45^. Furthermore, these studies focused on patients who either had SARS-CoV2 infection with a mild course or were hospitalized with a moderate-to-severe course. In addition, the previous analysis did not include the expected severity of COVID-19 through associations with established risk scores such as the ISARIC WHO 4C mortality score, another strength of our proposed study.

CVD is characterized by an augmented systemic inflammation triggering atherogenesis and - progression and is regulated by platelet and monocyte activation and the secretion of pro-inflammatory mediators, which play a crucial role in cell adhesion and migration ^23^. Consistent with and in addition to previous studies, we observed changes in the frequencies of innate immune cells like CD14^+^ monocytes, neutrophils, mature NK cells, and MAIT cells, as well as the adaptive immune cells CD4^+^CD8^+^, CD8^+^ naïve and central memory T cells, γδ T cells, early-like effector memory CD4^+^ T cells, and naïve B cell subsets in CVD patients compared to HD ^21, 25, 45–49^. Moreover, IL-6 levels were increased in CVD patients compared to HD. Thus, a chronic pro-inflammatory response in CVD patients might influence their immune signature compared to HD, leading to a more pronounced immune response to viral infections such as SARS-CoV-2.

We show that SARS-CoV-2-infected CVD patients show significant differences in immune cell populations’ distribution and phenotype compared to CVD patients without infections. Our data are consistent with other studies regarding, e.g., fewer non-classical monocytes in peripheral blood. These cells were shown to migrate to the site of organ injury, where they maintain the inflammatory processes within cardiac or pulmonary tissue ^40, 42, 44, 46^. As previously reported, we detected a more significant proportion of mature NK cells, CD14^+^CD16^+^ intermediate monocytes, activated populations of CD14^+^CD45RA^+^ monocytes, CD4^+^HLA-DR^+^CD38^+^ and CD8^+^ early T_em_ cells expressing HLA-DR^+^CD38^+^PD-1^+^ cells, as well as plasmablasts upon SARS-CoV-2 infection^4, 5, 8, 40, 42, 43^. In contrast, reduced numbers of ILCs, dendritic cell subsets cDC1 and cDC2, less activated/mature CD14^+^ monocytes, and CD8^+^ T cell subsets were observed in SARS-CoV-2 infected compared to uninfected CVD patients^4, 5, 8, 41, 42, 47^.

Furthermore, we describe phenotypical changes in SARS-CoV-2 infected CVD patients compared to uninfected CVD patients characterized by an overall higher expression of CD38 on plasmablasts, monocyte, NK cell, and T cell subsets ^40, 42, 43^ and lower expression of HLA-DR on DCs, CD14^+^ and CD14^+^CD16^+^ monocytes ^4, 40, 42, 46, 48^. Moreover, as described for NK cells ^4^, less CD16 was expressed on neutrophils, monocyte subsets, NKT cells, CD8^+,^ and CD4^+^ T cell subsets from CVD patients with severe COVID-19 compared to those with a mild infection. These data point towards reduced antibody-dependent cellular toxicity, thereby less killing of virus-infected cells (ADCC ^38^). In contrast to Georg et al., we did not detect CD16^+^ activated T cells in severely infected SARS-CoV-2 CVD patients ^49^. This may be due to differences in the cohorts (all-comers versus CVD patients) and medication before blood collection (Partial medication such as antibiotics or steroids compared to no medication in our study).

Consistent with another study, we found increased expression of the thrombin receptor CD141 on monocyte clusters from SARS-CoV-2 infected CVD patients ^42^. CD141 reduces blood coagulation which, among other mechanisms, may be an additional explanation for thromboembolic complications, especially in CVD patients and COVID-19 ^32, 42, 50^.

Plasmablasts and B cell subsets revealed a more inhibitory phenotype evidenced by reduced CD19 cell surface expression in SARS-CoV-2-infected CVD patients, whereas the latter was severity-dependent, indicating less B-cell receptor signaling^8, 37^.

In addition to other studies, we found that the remaining pDCs, regulating the type I interferon response ^31^, also showed a significantly more activated phenotype along with basophils and ILCs in SARS-CoV-2-infected CVD patients. pDCs and cDC subsets from SARS-CoV-2 infected CVD patients revealed a more migratory phenotype evidenced by their increased expression CCR7. In contrast, the chemokine receptor CXCR3, which is generally upregulated on activated NK and T cells^39^, was less expressed on neutrophils, mature NK cells, MAIT cells, and several CD4^+^ T cell subsets, implicating an impaired migratory capacity of these immune cells. In addition, the ligands for CXCR3, namely CXCL10, and CXCL11, were highly increased upon SARS-CoV-2 infection but also in several other viral infections and correlated with disease severity^51^. Thus one explanation for the reduced CXCR3 expression on many immune cells is downregulation upon ligand binding ^52^. Moreover, naïve CD4^+^ and CD8^+^ T cells from severely-infected CVD patients expressed higher levels of CD95 than those from mild-infected ones, indicating an impaired activation due to CD95-mediated inhibition of T-cell receptor signalling^53^. In summary, our data show an infection-induced activation of innate immune cells combined with reduced numbers and inhibitory phenotypes of DCs and adaptive immune cells in infected compared to uninfected CVD patients.

CVD patients with pre-existing cardiac and vascular dysfunction may benefit from intensified heart failure and anti-thrombotic therapy before progression to respiratory failure^4^. Since there is currently limited causal therapy for SARS-CoV-2 infection, early identification and treatment of prognostically relevant comorbidities are critical to prevent a fatal course triggered by the disease. We performed CombiROC analyses to predict the expected severity of COVID-19 at first patient contact based on a specific immune signature analyzed by flow cytometry. These analyses showed that a combination of only three immune cells detected in the blood at hospital admission (NKT cells, MAIT cells, and intermediate CD8^+^ effector T cells) is sufficient to distinguish CVD patients with mild from those with a severe course of infection.

Including characteristic findings in high-risk CVD patients, such as the specific immune signature, may improve the performance and generalizability of the 4C Mortality Score. This specific immune signature was independently associated with the severity of COVID-19 as determined by the ISARIC WHO 4C mortality score. In addition, there were no confounding factors in the OPLS-DA analysis, such as the comorbidities described above, suggesting that the immune signature may serve as an additional and objective tool for more intensive risk assessment of CVD patients with COVID-19 and increased risk of severe disease progression or infection-related disadvantages or complications, respectively. The ISARIC-WHO-4C mortality score is an easy-to-use, well-validated risk calculator for stratifying patients ^54^. However, the data collection of typical cardiovascular comorbidities such as hypertension, previous coronary artery disease, myocardial infarction, and stroke was not included in detail ^55^. Therefore, the risk assessment should be further improved for SARS-CoV-2 positive patients with pre-existing CVD, especially for very early stages of the viral infection. Here, we show that a pre-specified, objective analysis of the immune signature can identify CVD patients with an altered immune response at risk for severe courses of viral infections upon first admission or even at asymptomatic stages. These high-risk CVD patients could benefit from intensified monitoring and early anti-inflammatory treatment strategies, thereby preventing long-term ICU treatment or fatal outcomes. Another advantage of the immune signature is good discrimination between mild and severe courses of COVID-19 by an objective laboratory method that is not dependent on the sometimes hard-to-assess, sometimes poorly-defined, clinical parameters associated with unfavorable outcomes.

Nevertheless, the study has some limitations. Although we have studied a relatively large cohort of CVD patients with and without SARS-CoV-2 infection and HD, the number of patients with severe courses is limited. We analyzed circulating immune cells and their phenotype, more precise PBMCs. Therefore, we lose a large part of the neutrophil granulocytes in the analysis, shown to be crucial for the distinction between the fatal and non-fatal outcomes of the disease ^40^. And while knowledge of peripheral immune cells is essential for understanding pathogenic and protective immune responses to SARS-CoV-2 infection, it does not cover the immune response at the infection site.

In conclusion, our study revealed that a specific immune signature is associated with the severity of COVID-19 in patients with CVD and can predict the course of the disease already upon first admission to the hospital. The early determination of the immune signature might enable the treating clinicians to provide the best possible pharmacological and device-based care upon hospitalization and improve clinical outcomes.

## Data Availability

All data produced in the present study are available upon reasonable request to the authors.

## Acknowledgment

We thank the Core Facility Flow Cytometry of the Medical Faculty Tübingen for technical help.

## Author contributions

MG performed experimental analysis, data acquisition, and preparation of Figures. CP and SG performed the Legendplex analysis. MS and TH analyzed data using R. KALM, and MPG were involved in the conception and design of the study. KALM was responsible for the patient cohort and their classification and manuscript writing. SEA was responsible for the concept and study design, data analysis, data interpretation, drafting of the manuscript, preparation of Figures, and revising it critically for important intellectual content.

## Data Availability Statement

For original data, please contact Stella E. Autenrieth at.

## References

1. Koutsakos M, Kedzierska K. A race to determine what drives COVID-19 severity. Nature. 2020;583(7816):366–368. doi:10.1038/d41586-020-01915-3

2. Zhang X, Tan Y, Ling Y, Lu G, Liu F, Yi Z, Jia X, Wu M, Shi B, Xu S, Chen J, Wang W, Chen B, Jiang L, Yu S, Lu J, Wang J, Xu M, Yuan Z, Zhang Q, Zhang X, Zhao G, Wang S, Chen S, Lu H. Viral and host factors related to the clinical outcome of COVID-19. Nature. 2020;583(7816):437–440. doi:10.1038/s41586-020-2355-0

3. Thevarajan I, Nguyen THO, Koutsakos M, Druce J, Caly L, Sandt CE van de, Jia X, Nicholson S, Catton M, Cowie B, Tong SYC, Lewin SR, Kedzierska K. Breadth of concomitant immune responses prior to patient recovery: a case report of non-severe COVID-19. Nat Med. 2020;26(4):453–455. doi:10.1038/s41591-020-0819-2

4. Kuri-Cervantes L, Pampena MB, Meng W, Rosenfeld AM, Ittner CAG, Weisman AR, Agyekum RS, Mathew D, Baxter AE, Vella LA, Kuthuru O, Apostolidis SA, Bershaw L, Dougherty J, Greenplate AR, Pattekar A, Kim J, Han N, Gouma S, Weirick ME, Arevalo CP, Bolton MJ, Goodwin EC, Anderson EM, Hensley SE, Jones TK, Mangalmurti NS, Prak ETL, Wherry EJ, Meyer NJ, Betts MR. Comprehensive mapping of immune perturbations associated with severe COVID-19. Sci Immunol. 2020;5(49):eabd7114. doi:10.1126/sciimmunol.abd7114

5. Mathew D, Giles JR, Baxter AE, Oldridge DA, Greenplate AR, Wu JE, Alanio C, Kuri-Cervantes L, Pampena MB, D’Andrea K, Manne S, Chen Z, Huang YJ, Reilly JP, Weisman AR, Ittner CAG, Kuthuru O, Dougherty J, Nzingha K, Han N, Kim J, Pattekar A, Goodwin EC, Anderson EM, Weirick ME, Gouma S, Arevalo CP, Bolton MJ, Chen F, Lacey SF, Ramage H, Cherry S, Hensley SE, Apostolidis SA, Huang AC, Vella LA, Unit TUpCP, Betts MR, Meyer NJ, Wherry EJ, Alam Z, Addison MM, Byrne KT, Chandra A, Descamps HC, Kaminskiy Y, Hamilton JT, Noll JH, Omran DK, Perkey E, Prager EM, Pueschl D, Shah JB, Shilan JS, Vanderbeck AN. Deep immune profiling of COVID-19 patients reveals distinct immunotypes with therapeutic implications. Science. 2020;369(6508):eabc8511. doi:10.1126/science.abc8511

6. Long QX, Tang XJ, Shi QL, Li Q, Deng HJ, Yuan J, Hu JL, Xu W, Zhang Y, Lv FJ, Su K, Zhang F, Gong J, Wu B, Liu XM, Li JJ, Qiu JF, Chen J, Huang AL. Clinical and immunological assessment of asymptomatic SARS-CoV-2 infections. Nat Med. 2020;26(8):1200–1204. doi:10.1038/s41591-020-0965-6

7. Lucas C, Wong P, Klein J, Castro TBR, Silva J, Sundaram M, Ellingson MK, Mao T, Oh JE, Israelow B, Takahashi T, Tokuyama M, Lu P, Venkataraman A, Park A, Mohanty S, Wang H, Wyllie AL, Vogels CBF, Earnest R, Lapidus S, Ott IM, Moore AJ, Muenker MC, Fournier JB, Campbell M, Odio CD, Casanovas-Massana A, Obaid A, Lu-Culligan A, Nelson A, Brito A, Nunez A, Martin A, Watkins A, Geng B, Kalinich C, Harden C, Todeasa C, Jensen C, Kim D, McDonald D, Shepard D, Courchaine E, White EB, Song E, Silva E, Kudo E, DeIuliis G, Rahming H, Park HJ, Matos I, Nouws J, Valdez J, Fauver J, Lim J, Rose KA, Anastasio K, Brower K, Glick L, Sharma L, Sewanan L, Knaggs L, Minasyan M, Batsu M, Petrone M, Kuang M, Nakahata M, Campbell M, Linehan M, Askenase MH, Simonov M, Smolgovsky M, Sonnert N, Naushad N, Vijayakumar P, Martinello R, Datta R, Handoko R, Bermejo S, Prophet S, Bickerton S, Velazquez S, Alpert T, Rice T, Khoury-Hanold W, Peng X, Yang Y, Cao Y, Strong Y, Herbst R, Shaw AC, Medzhitov R, Schulz WL, Grubaugh ND, Cruz CD, Farhadian S, Ko AI, et al. Longitudinal analyses reveal immunological misfiring in severe COVID-19. Nature. 2020;584(7821):463–469. doi:10.1038/s41586-020-2588-y

8. Laing AG, Lorenc A, Barrio I del M del, Das A, Fish M, Monin L, Muñoz-Ruiz M, McKenzie DR, Hayday TS, Francos-Quijorna I, Kamdar S, Joseph M, Davies D, Davis R, Jennings A, Zlatareva I, Vantourout P, Wu Y, Sofra V, Cano F, Greco M, Theodoridis E, Freedman JD, Gee S, Chan JNE, Ryan S, Bugallo-Blanco E, Peterson P, Kisand K, Haljasmägi L, Chadli L, Moingeon P, Martinez L, Merrick B, Bisnauthsing K, Brooks K, Ibrahim MAA, Mason J, Gomez FL, Babalola K, Abdul-Jawad S, Cason J, Mant C, Seow J, Graham C, Doores KJ, Rosa FD, Edgeworth J, Shankar-Hari M, Hayday AC. A dynamic COVID-19 immune signature includes associations with poor prognosis. Nat Med. 2020;26(10):1623–1635. doi:10.1038/s41591-020-1038-6

9. MD PCH, MD YW, MD PXL, PhD PLR, MD PJZ, MD YH, MD PLZ, MS GF, MDc JX, PhD XG, MD PZC, MD TY, MD JX, MD YW, MD PWW, MD PXX, MD WY, MD HL, MD ML, MS YX, PhD PHG, PhD PLG, MD PJX, MD PGW, MD PRJ, MD PZG, PhD QJ, PhD PJW, MD PBC. Clinical features of patients infected with 2019 novel coronavirus in Wuhan, China. The Lancet. 2020;395(10223):497–506. doi:10.1016/s0140-6736(20)30183-5

10. Zhou F, Yu T, Du R, Fan G, Liu Y, Liu Z, Xiang J, Wang Y, Song B, Gu X, Guan L, Wei Y, Li H, Wu X, Xu J, Tu S, Zhang Y, Chen H, Cao B. Clinical course and risk factors for mortality of adult inpatients with COVID-19 in Wuhan, China: a retrospective cohort study. The Lancet. 2020;395(10229):1054–1062. doi:10.1016/s0140-6736(20)30566-3

11. Jose RJ, Manuel A. COVID-19 cytokine storm: the interplay between inflammation and coagulation. The Lancet Respiratory. Published online April 24, 2020:1–2. doi:10.1016/s2213-2600(20)30216-2

12. Mangalmurti N, Hunter CA. Cytokine Storms: Understanding COVID-19. Immunity. 2020;53(1):19–25. doi:10.1016/j.immuni.2020.06.017

13. Cardiology ES of. ESC Guidance for the Diagnosis and Management of CV Disease during the COVID-19 Pandemic. Published online April 28, 2020:1–119.

14. Oren O, Yang EH, Molina JR, Bailey KR, Blumenthal RS, Kopecky SL. Cardiovascular Health and Outcomes in Cancer Patients Receiving Immune Checkpoint Inhibitors. The American journal of cardiology. Published online March 5, 2020. doi:10.1016/j.amjcard.2020.02.016

15. Xiong TY, Redwood S, Prendergast B, Chen M. Coronaviruses and the cardiovascular system: acute and long-term implications. Eur Heart J. 2020;41(19):1798–1800. doi:10.1093/eurheartj/ehaa231

16. Chen C, Zhou Y, Wang DW. SARS-CoV-2: a potential novel etiology of fulminant myocarditis. Herz. 2020;45(3):230–232. doi:10.1007/s00059-020-04909-z

17. Shi S, Qin M, Shen B, Cai Y, Liu T, Yang F, Gong W, Liu X, Liang J, Zhao Q, Huang H, Yang B, Huang C. Association of Cardiac Injury With Mortality in Hospitalized Patients With COVID-19 in Wuhan, China. Jama Cardiol. 2020;5(7):802–810. doi:10.1001/jamacardio.2020.0950

18. Tay JY, Lim PL, Marimuthu K, Sadarangani SP, Ling LM, Ang BSP, Chan M, Leo YS, Vasoo S. De-isolating COVID-19 Suspect Cases: A Continuing Challenge. Clin Infect Dis. 2020;71(15):ciaa179. doi:10.1093/cid/ciaa179

19. Tang N, Bai H, Chen X, Gong J, Li D, Sun Z. Anticoagulant treatment is associated with decreased mortality in severe coronavirus disease 2019 patients with coagulopathy. J Thromb Haemost. 2020;18(5):1094–1099. doi:10.1111/jth.14817

20. Wolf D, Zirlik A, Ley K. Beyond vascular inflammation—recent advances in understanding atherosclerosis. Cell Mol Life Sci. 2015;72(20):3853–3869. doi:10.1007/s00018-015-1971-6

21. Wolf D, Ley K. Immunity and Inflammation in Atherosclerosis. Circ Res. 2019;124(2):315–327. doi:10.1161/circresaha.118.313591

22. Bahrar H, Bekkering S, Stienstra R, Netea MG, Riksen NP. Innate immune memory in cardiometabolic disease. Cardiovasc Res. Published online 2023. doi:10.1093/cvr/cvad030

23. Swirski FK, Nahrendorf M. Cardioimmunology: the immune system in cardiac homeostasis and disease. Nature. 2018;18(12):1–12. doi:10.1038/s41577-018-0065-8

24. Woollard KJ, Geissmann F. Monocytes in atherosclerosis: subsets and functions. Nature reviews Cardiology. 2010;7(2):77–86. doi:10.1038/nrcardio.2009.228

25. Roy P, Orecchioni M, Ley K. How the immune system shapes atherosclerosis: roles of innate and adaptive immunity. Nat Rev Immunol. 2022;22(4):251–265. doi:10.1038/s41577-021-00584-1

26. McInnes L, Healy J, Melville J. UMAP: Uniform Manifold Approximation and Projection for Dimension Reduction. Arxiv. Published online 2018.

27. Gassen SV, Callebaut B, Helden MJV, Lambrecht BN, Demeester P, Dhaene T, Saeys Y. FlowSOM: Using self-organizing maps for visualization and interpretation of cytometry data. Brinkman RR, Aghaeepour Nima, Finak Greg, Gottardo Raphael, Mosmann Tim, Scheuermann RH, eds. Cytometry Part B: Clinical Cytometry. 2015;87(7):636–645. doi:10.1002/cyto.a.22625

28. Mazzara S, Rossi RL, Grifantini R, Donizetti S, Abrignani S, Bombaci M. CombiROC: an interactive web tool for selecting accurate marker combinations of omics data. Sci Rep-uk. 2017;7(1):45477. doi:10.1038/srep45477

29. Bombaci M, Rossi RL. Proteomics for Biomarker Discovery, Methods and Protocols. Methods Mol Biology. 2019;1959:247–259. doi:10.1007/978-1-4939-9164-8_16

30. Park LM, Lannigan J, Jaimes MC. OMIP-069: Forty-Color Full Spectrum Flow Cytometry Panel for Deep Immunophenotyping of Major Cell Subsets in Human Peripheral Blood. Cytom Part A. 2020;97(10):1044–1051. doi:10.1002/cyto.a.24213

31. Reizis B. Plasmacytoid Dendritic Cells: Development, Regulation, and Function. Immunity. 2019;50(1):37–50. doi:10.1016/j.immuni.2018.12.027

32. Ferrari F, Martins VM, Teixeira M, Santos RD, Stein R. COVID-19 and Thromboinflammation: Is There a Role for Statins? Clinics. 2021;76:e2518. doi:10.6061/clinics/2021/e2518

33. Blankenberg S, Tiret L, Bickel C, Peetz D, Cambien F, Meyer J, Rupprecht HJ, Investigators A. Interleukin-18 Is a Strong Predictor of Cardiovascular Death in Stable and Unstable Angina. Circulation. 2002;106(1):24–30. doi:10.1161/01.cir.0000020546.30940.92

34. Pearson TA, Mensah GA, Alexander RW, Anderson JL, Cannon RO, Criqui M, Fadl YY, Fortmann SP, Hong Y, Myers GL, Rifai N, Smith SC, Taubert K, Tracy RP, Vinicor F, Prevention C for DC and, Association AH. Markers of Inflammation and Cardiovascular Disease. Circulation. 2003;107(3):499–511. doi:10.1161/01.cir.0000052939.59093.45

35. Imai T, Baba M, Nishimura M, Kakizaki M, Takagi S, Yoshie O. The T Cell-directed CC Chemokine TARC Is a Highly Specific Biological Ligand for CC Chemokine Receptor 4*. J Biol Chem. 1997;272(23):15036–15042. doi:10.1074/jbc.272.23.15036

36. Hinks TSC, Zhang XW. MAIT Cell Activation and Functions. Front Immunol. 2020;11:1014. doi:10.3389/fimmu.2020.01014

37. Wang K, Wei G, Liu D. CD19: a biomarker for B cell development, lymphoma diagnosis and therapy. Exp Hematology Oncol. 2012;1(1):36. doi:10.1186/2162-3619-1-36

38. Yeap WH, Wong KL, Shimasaki N, Teo ECY, Quek JKS, Yong HX, Diong CP, Bertoletti A, Linn YC, Wong SC. CD16 is indispensable for antibody-dependent cellular cytotoxicity by human monocytes. Sci Rep-uk. 2016;6(1):34310. doi:10.1038/srep34310

39. Wang X, Zhang Y, Wang S, Ni H, Zhao P, Chen G, Xu B, Yuan L. The role of CXCR3 and its ligands in cancer. Frontiers Oncol. 2022;12:1022688. doi:10.3389/fonc.2022.1022688

40. Wilk AJ, Lee MJ, Wei B, Parks B, Pi R, Martínez-Colón GJ, Ranganath T, Zhao NQ, Taylor S, Becker W, Biobank SC 19, Ranganath T, Zhao NQ, Wilk AJ, Vergara R, McKechnie JL, Parte L de la, Dantzler KW, Ty M, Kathale N, Martinez-Colon GJ, Rustagi A, Ivison G, Pi R, Lee MJ, Brewer R, Hollis T, Baird A, Ugur M, Tal M, Bogusch D, Nahass G, Haider K, Tran KQT, Simpson L, Din H, Roque J, Mann R, Chang I, Do E, Fernandes A, Lyu SC, Zhang W, Manohar M, Krempski J, Visweswaran A, Zudock EJ, Jee K, Kumar K, Newberry JA, Quinn JV, Schreiber D, Ashley EA, Blish CA, Blomkalns AL, Nadeau KC, O’Hara R, Rogers AJ, Yang S, Jimenez-Morales D, Blomkalns AL, O’Hara R, Ashley EA, Nadeau KC, Yang S, Holmes S, Rabinovitch M, Rogers AJ, Greenleaf WJ, Blish CA. Multi-omic profiling reveals widespread dysregulation of innate immunity and hematopoiesis in COVID-19. J Exp Med. 2021;218(8):e20210582. doi:10.1084/jem.20210582

41. Breton G, Mendoza P, Hägglöf T, Oliveira TY, Schaefer-Babajew D, Gaebler C, Turroja M, Hurley A, Caskey M, Nussenzweig MC. Persistent cellular immunity to SARS-CoV-2 infection. J Exp Med. 2021;218(4). doi:10.1084/jem.20202515

42. Kvedaraite E, Hertwig L, Sinha I, Ponzetta A, Myrberg IH, Lourda M, Dzidic M, Akber M, Klingström J, Folkesson E, Muvva JR, Chen P, Gredmark-Russ S, Brighenti S, Norrby-Teglund A, Eriksson LI, Rooyackers O, Aleman S, Strålin K, Ljunggren HG, Ginhoux F, Björkström NK, Henter JI, Svensson M, Group KKC 19 S, Sandberg JT, Bergsten H, Björkström NK, Brighenti S, Buggert M, Butrym M, Chambers BJ, Chen P, Cornillet M, Cuapio A, Lozano ID, Dzidic M, Emgård J, Flodström-Tullberg M, Gorin JB, Gredmark-Russ S, Haroun-Izquierdo A, Hertwig L, Kalsum S, Klingström J, Kokkinou E, Kvedaraite E, Ljunggren HG, Marquardt N, Lourda M, Maleki KT, Malmberg KJ, Michaëlsson J, Mjösberg J, Moll K, Muvva JR, Norrby-Teglund A, Medina LMP, Parrot T, Radler L, Ringqvist E, Sandberg JK, Sekine T, Soini T, Svensson M, Tynell J, Kries A von, Wullimann D, Perez-Potti A, Rivera-Ballesteros O, Maucourant C, Varnaite R, Akber M, Berglin L, Brownlie D, Loreti MG, Sohlberg E, Kammann T, Henriksson E, Strålin K, Aleman S, Sönnerborg A, Dillner L, Färnert A, Glans H, Nauclér P, Rooyackers O, Mårtensson J, Eriksson LI, Persson BP, Grip J, Unge C. Major alterations in the mononuclear phagocyte landscape associated with COVID-19 severity. P Natl Acad Sci Usa. 2021;118(6):e2018587118. doi:10.1073/pnas.2018587118

43. Koutsakos M, Rowntree LC, Hensen L, Chua BY, Sandt CE van de, Habel JR, Zhang W, Jia X, Kedzierski L, Ashhurst TM, Putri GH, Marsh-Wakefield F, Read MN, Edwards DN, Clemens EB, Wong CY, Mordant FL, Juno JA, Amanat F, Audsley J, Holmes NE, Gordon CL, Smibert OC, Trubiano JA, Hughes CM, Catton M, Denholm JT, Tong SYC, Doolan DL, Kotsimbos TC, Jackson DC, Krammer F, Godfrey DI, Chung AW, King NJC, Lewin SR, Wheatley AK, Kent SJ, Subbarao K, McMahon J, Thevarajan I, Nguyen THO, Cheng AC, Kedzierska K. Integrated immune dynamics define correlates of COVID-19 severity and antibody responses. Cell Reports Medicine. 2021;2(3):100208. doi:10.1016/j.xcrm.2021.100208

44. Gatti A, Radrizzani D, Viganò P, Mazzone A, Brando B. Decrease of Non-Classical and Intermediate Monocyte Subsets in Severe Acute SARS-CoV-2 Infection. Cytom Part A. 2020;97(9):887–890. doi:10.1002/cyto.a.24188

45. Mueller YM, Schrama TJ, Ruijten R, Schreurs MWJ, Grashof DGB, Werken HJG van de, Lasinio GJ, Álvarez-Sierra D, Kiernan CH, Eiro MDC, Meurs M van, Brouwers-Haspels I, Zhao M, Li L, Wit H de, Ouzounis CA, Wilmsen MEP, Alofs TM, Laport DA, Wees T van, Kraker G, Jaimes MC, Bockstael SV, Hernández-González M, Rokx C, Rijnders BJA, Pujol-Borrell R, Katsikis PD. Stratification of hospitalized COVID-19 patients into clinical severity progression groups by immuno-phenotyping and machine learning. Nat Commun. 2022;13(1):915. doi:10.1038/s41467-022-28621-0

46. Mueller KAL, Langnau C, Günter M, Pöschel S, Gekeler S, Petersen-Uribe Á, Kreisselmeier KP, Klingel K, Bösmüller H, Li B, Jaeger P, Castor T, Rath D, Gawaz MP, Autenrieth SE. Numbers and phenotype of non-classical CD14dimCD16+ monocytes are predictors of adverse clinical outcome in patients with coronary artery disease and severe SARS-CoV-2 infection. Cardiovasc Res. 2021;117(1):(1):224–239. doi:10.1093/cvr/cvaa328

47. Winheim E, Rinke L, Lutz K, Reischer A, Leutbecher A, Wolfram L, Rausch L, Kranich J, Wratil PR, Huber JE, Baumjohann D, Rothenfusser S, Schubert B, Hilgendorff A, Hellmuth JC, Scherer C, Muenchhoff M, Bergwelt-Baildon M von, Stark K, Straub T, Brocker T, Keppler OT, Subklewe M, Krug AB. Impaired function and delayed regeneration of dendritic cells in COVID-19. Plos Pathog. 2021;17(10):e1009742. doi:10.1371/journal.ppat.1009742

48. Zhou R, To KKW, Wong YC, Liu L, Zhou B, Li X, Huang H, Mo Y, Luk TY, Lau TTK, Yeung P, Chan WM, Wu AKL, Lung KC, Tsang OTY, Leung WS, Hung IFN, Yuen KY, Chen Z. Acute SARS-CoV-2 infection impairs dendritic cell and T cell responses. Immunity. 2020;53(4):864–877.e5. doi:10.1016/j.immuni.2020.07.026

49. Georg P, Astaburuaga-García R, Bonaguro L, Brumhard S, Michalick L, Lippert LJ, Kostevc T, Gäbel C, Schneider M, Streitz M, Demichev V, Gemünd I, Barone M, Tober-Lau P, Helbig ET, Hillus D, Petrov L, Stein J, Dey HP, Paclik D, Iwert C, Mülleder M, Aulakh SK, Djudjaj S, Bülow RD, Mei HE, Schulz AR, Thiel A, Hippenstiel S, Saliba AE, Eils R, Lehmann I, Mall MA, Stricker S, Röhmel J, Corman VM, Beule D, Wyler E, Landthaler M, Obermayer B, Stillfried S von, Boor P, Demir M, Wesselmann H, Suttorp N, Uhrig A, Müller-Redetzky H, Nattermann J, Kuebler WM, Meisel C, Ralser M, Schultze JL, Aschenbrenner AC, Thibeault C, Kurth F, Sander LE, Blüthgen N, Sawitzki B, Group PC 19 S. Complement activation induces excessive T cell cytotoxicity in severe COVID-19. Cell. 2022;185(3):493–512.e25. doi:10.1016/j.cell.2021.12.040

50. Mizurini DM, Hottz ED, Bozza PT, Monteiro RQ. Fundamentals in Covid-19-Associated Thrombosis: Molecular and Cellular Aspects. Frontiers Cardiovasc Medicine. 2021;8:785738. doi:10.3389/fcvm.2021.785738

51. Khalil BA, Elemam NM, Maghazachi AA. Chemokines and chemokine receptors during COVID-19 infection. Comput Struct Biotechnology J. 2021;19:976–988. doi:10.1016/j.csbj.2021.01.034

52. Meiser A, Mueller A, Wise EL, McDonagh EM, Petit SJ, Saran N, Clark PC, Williams TJ, Pease JE. The Chemokine Receptor CXCR3 Is Degraded following Internalization and Is Replenished at the Cell Surface by De Novo Synthesis of Receptor. J Immunol. 2008;180(10):6713–6724. doi:10.4049/jimmunol.180.10.6713

53. Strauss G, Lindquist JA, Arhel N, Felder E, Karl S, Haas TL, Fulda S, Walczak H, Kirchhoff F, Debatin KM. CD95 co-stimulation blocks activation of naive T cells by inhibiting T cell receptor signaling. J Exp Med. 2009;206(6):1379–1393. doi:10.1084/jem.20082363

54. Knight SR, Ho A, Pius R, Buchan I, Carson G, Drake TM, Dunning J, Fairfield CJ, Gamble C, Green CA, Gupta R, Halpin S, Hardwick HE, Holden KA, Horby PW, Jackson C, Mclean KA, Merson L, Nguyen-Van-Tam JS, Norman L, Noursadeghi M, Olliaro PL, Pritchard MG, Russell CD, Shaw CA, Sheikh A, Solomon T, Sudlow C, Swann OV, Turtle LC, Openshaw PJ, Baillie JK, Semple MG, Docherty AB, Harrison EM, Baillie JK, Semple MG, Openshaw PJ, Carson G, Alex B, Bach B, Barclay WS, Bogaert D, Chand M, Cooke GS, Docherty AB, Dunning J, Filipe A da S, Fletcher T, Green CA, Harrison EM, Hiscox JA, Ho AYW, Horby PW, Ijaz S, Khoo S, Klenerman P, Law A, Lim WS, Mentzer AJ, Merson L, Meynert AM, Noursadeghi M, Moore SC, Palmarini M, Paxton WA, Pollakis G, Price N, Rambaut A, Robertson DL, Russell CD, Sancho-Shimizu V, Scott JT, Sigfrid L, Solomon T, Sriskandan S, Stuart D, Summers C, Tedder RS, Thomson EC, Thwaites RS, Turtle LC, Zambon M, Hardwick H, Donohue C, Ewins J, Oosthuyzen W, Griffiths F, Norman L, Pius R, Drake TM, Fairfield CJ, Knight S, Mclean KA, Murphy D, Shaw CA, Dalton J, Girvan M, et al. Risk stratification of patients admitted to hospital with covid-19 using the ISARIC WHO Clinical Characterisation Protocol: development and validation of the 4C Mortality Score. Bmj. 2020;370:m3339. doi:10.1136/bmj.m3339

55. Guan WJ, Liang WH, Zhao Y, Liang HR, Chen ZS, Li YM, Liu XQ, Chen RC, Tang CL, Wang T, Ou CQ, Li L, Chen PY, Sang L, Wang W, Li JF, Li CC, Ou LM, Cheng B, Xiong S, Ni ZY, Xiang J, Hu Y, Liu L, Shan H, Lei CL, Peng YX, Wei L, Liu Y, Hu YH, Peng P, Wang JM, Liu JY, Chen Z, Li G, Zheng ZJ, Qiu SQ, Luo J, Ye CJ, Zhu SY, Cheng LL, Ye F, Li SY, Zheng JP, Zhang NF, Zhong NS, He JX, COVID-19 CMTEG for. Comorbidity and its impact on 1590 patients with COVID-19 in China: a nationwide analysis. Eur Respir J. 2020;55(5):2000547. doi:10.1183/13993003.00547-2020

