## Supplemental Material for "Immune signature of patients with cardiovascular disease – in-depth immunophenotyping predicts increased risk for a severe course of COVID-19"

| Approx. Emission Wavelength (nm) | UV | Violet | Blue | Yellow Green | Red |
|---|---|---|---|---|---|
| 395 | CD45RA BUV395 | | | | |
| 420 | Viability UV | CCR7 BV421 | | | |
| 440 | | CD123 SB436 | | | |
| 450 | | CD161 eFluor450 | | | |
| 480 | | IgD BV480 | | | |
| 500 | CD16 BUV496 | CD3 BV510 | CD141 BB515 | | |
| 520 | | | CD57 FITC | | |
| 550 | | CD20 Pacific Orange | CD14 Spark Blue 550 | | |
| 570 | CCR5 BUV563 | IgM BV570 | | CD25 PE | |
| 580 | | | | CD4 CF568 | |
| 600 | | IgG BV605 | | CD24 PE Dazzle594 | |
| 660 | CD11c BUV661 | CD28 BV650 | | | CD27 APC |
| 680 | | | CD45 PerCP | CD95 PE Cy5 | CD1c AlexaFluor 647 |
| 690 | | | CD11b PerCP Cy5.5 | | CD19 Spark NIR 685 |
| 700 | | CCR6 BV711 | TCR $\gamma\delta$ PerCP eFluor710 | | CD127 APC-R700 |
| 730 | CD56 BUV737 | | | | |
| 750 | | CXCR5 BV750 | | | |
| 780 | | PD-1 BV785 | | CXCR3 PE Cy7 | HLA DR APC Fire750 |
| 800 | CD8 BUV805 | | | | CD38 APC Fire 810 |

T cells
NK cells
B cells
Monocytes
DCs

**Supplemental Figure 1: 36-color Panel used for the analysis of immune cell populations**
Antibody panel used for immune cell phenotyping on a five laser Aurora spectral flow cytometer. Colors depict the marker expression on the indicated cell populations.

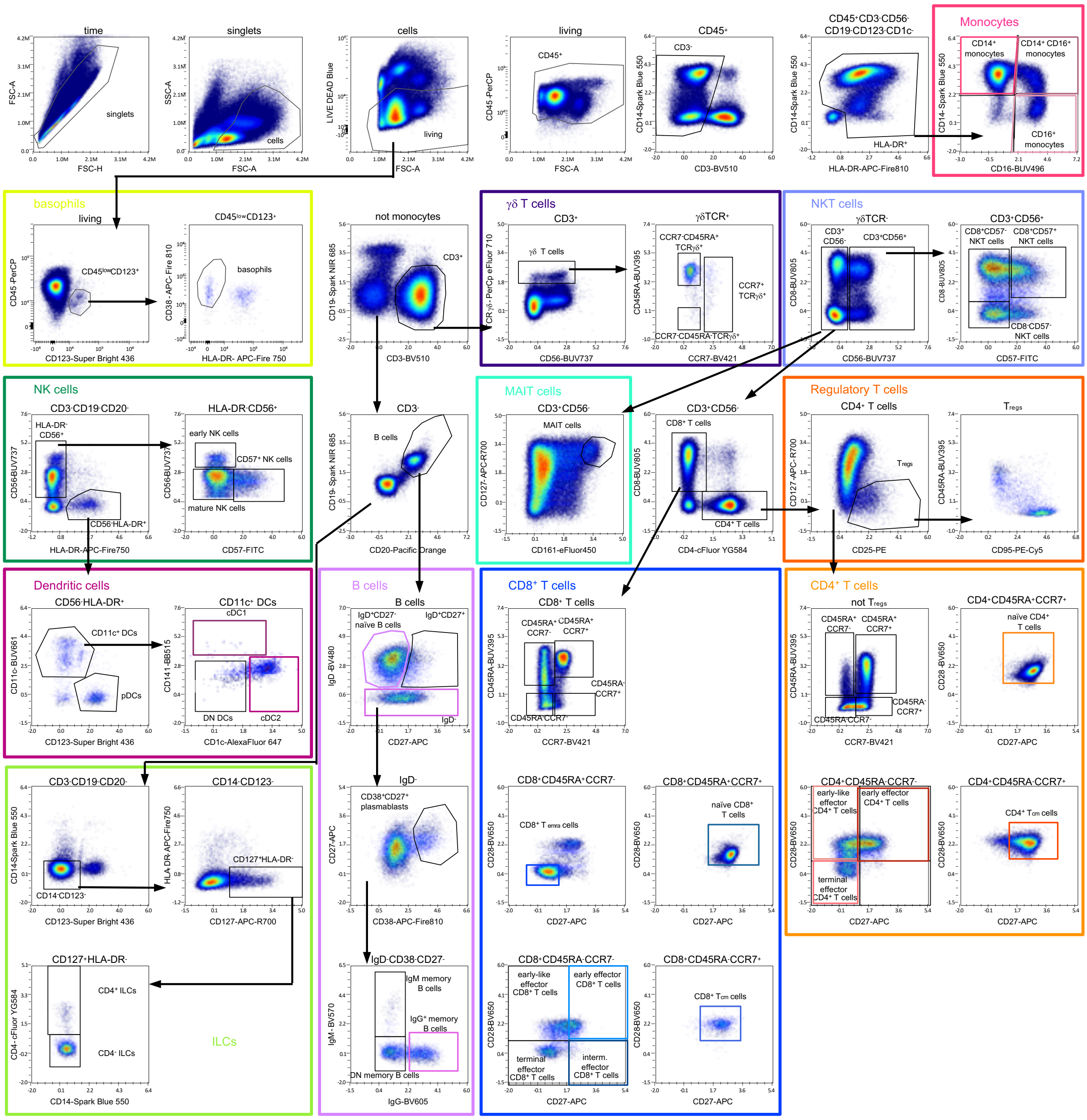


**Supplemental Figure 2: Manual gating strategy used to identify the main cellular subsets**. The arrows describe the relationship of the cells across the plots. FlowAI was used to clean the flow cytometry data from anomalies in flow rate, signal acquisition, and dynamic range (Monaco, G. *et al.* flowAI: automatic and interactive anomaly discerning tools for flow cytometry data. *Bioinformatics* 32, 2473–2480 (2016)). Thereafter doublets, debris, and dead cells were excluded. Basophils (yellow) were marked as $CD45^{low}CD123^{+}HLA\text{-}DR^{-}CD38^{+}$. Leukocytes were defined as $CD45^{+}$. Monocytes were then classified by the exclusion of lymphocyte markers as $CD45^{+}CD3^{-}CD56^{-}CD19^{-}CD123^{-}CD1c^{-}$ followed by CD14 and CD16 expression as classical ($CD14^{+}CD16^{-}$), intermediate ($CD14^{+}CD16^{+}$), and non-classical ($CD14^{-}CD16^{+}$). Subsequently, monocytes were excluded from further analysis of $CD45^{+}$ cells (not-gate). The further gating strategy is often based on the use of not-gates. Next, via the expression of CD3 γδ T cells (purple) and NKT cells (lilac) were defined as $CD3^{+}TCR\ \gamma\delta^{+}$ subsets and $CD3^{+}CD56^{+}$ subsets, respectively. $CD3^{+}CD56^{-}$ cells were further separated into $CD4^{+}$, $CD8^{+}$, $CD4^{+}CD8^{+}$ T cells. Regulatory T cells ($T_{regs}$) were identified from the $CD4^{+}$ population as $CD127^{low/-}CD25^{hi}$ cells, and CD95 and CD45RA expressions were used to classify these cells (red) further. CCR7, CD45RA, CD27, and CD28 expressions were used for in-depth characterization of memory/effector CD4 and CD8 T cell subsets (orange and blue, respectively). $CD3^{-}CD19^{+}CD20^{+}$ cells were defined as B cells (violet) and further gated as $IgD^{+}CD27^{-}$ naïve B cells, $IgD^{+}CD27^{+}$ B cells, or $IgD^{-}CD27^{+}CD38^{+}$ plasmablasts or $IgD^{-}$ memory B cells based on IgG or IgM expression. $CD3^{-}CD19^{-}CD20^{-}$ cells were further defined as $HLA\text{-}DR^{-}CD56^{+}$ NK cells with early NK ($CD56^{hi}CD57^{-}$), mature NK ($CD56^{+}CD57^{-}$), and terminal NK ($CD56^{+}CD57^{+}$) cell subsets (green) or $CD123^{-}CD14^{-}HLA\text{-}DR^{-}CD127^{+}$ ILCs (light green). The latter can be further characterized by CD4 expression. Dendritic cells (DCs, pink) were identified first by gating on $CD3^{-}CD19^{-}CD20^{-}CD56^{-}HLA\text{-}DR^{+}$, and from there, $CD123^{+}$ (pDCs) and $CD11c^{+}$ DCs were identified. $CD11c^{+}$ DCs were further divided into CD141 expressing cDC1s and CD1c expressing cDC2s. All data presented is derived from frozen PBMCs of one healthy donor.

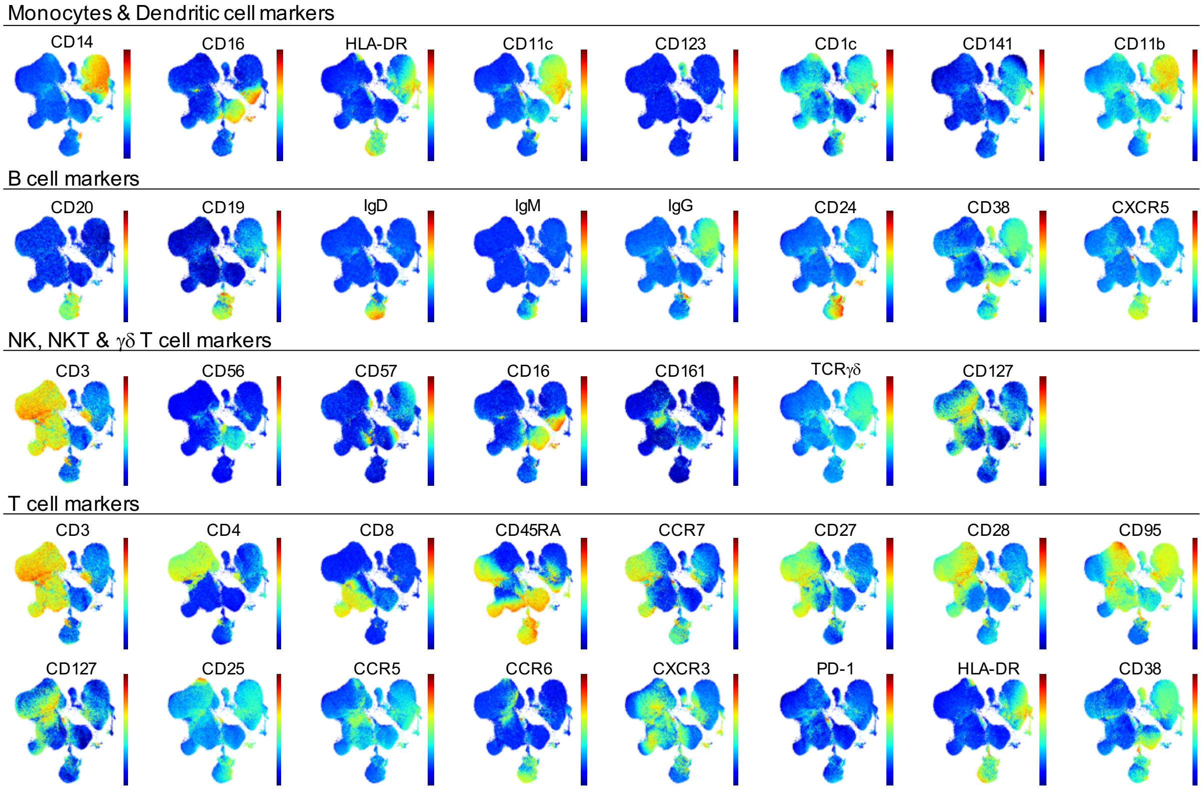


**Supplemental Figure 3: UMAP Dimensional Data Reduction of 36-color Panel**
Visualization of the expression of phenotypic markers on PBMCs from healthy donors using UMAP. Marker expression intensity is indicated by the scale bar to the right of each plot, where red is high and blue is low.

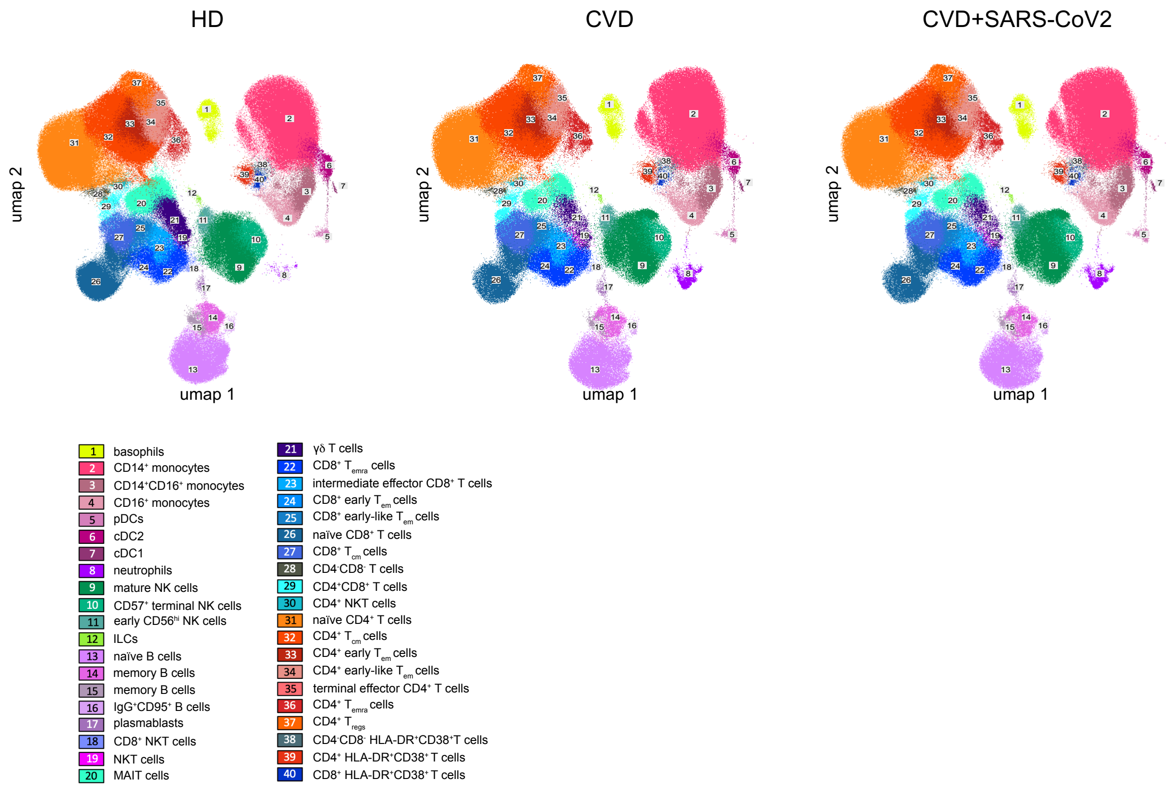


**Supplemental Figure 4:** High-dimensional data analysis of PBMCs displaying 40 FlowSOM clusters projected onto two UMAP dimensions. The UMAP-plots show concatenated events from PBMC samples from HD (37), CVD (20), and CVD + SARS-CoV-2-infected (37).

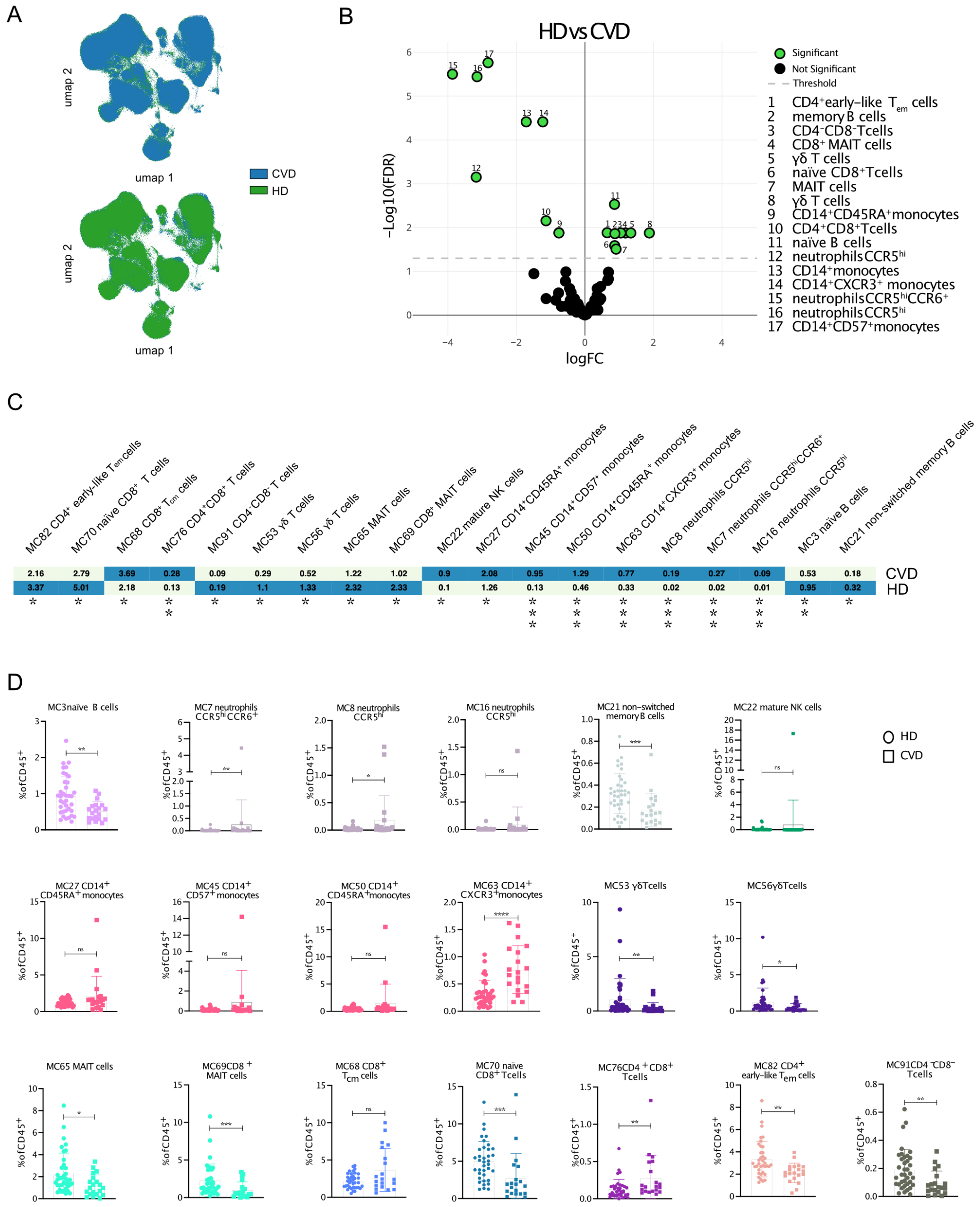


**Supplemental Figure 5: Differences between immune cell populations in CVD patients and HD**
**A** The overlay UMAP-plot shows concatenated events from HD (37) and CVD patient (20) samples. **B** Volcano plot of statistical analysis comparing the frequency of cells in the defined FSOM clusters (Figure 1c) from CVD patients and HD using edgeR. Significant different clusters are depicted in green. **C** Clustered heatmap displaying the frequency of cells in the defined FSOM clusters from 36 concatenated samples from HD (lower row) and 16 CVD patients (upper row). The frequency is displayed per column on a scale from white (low) to blue (high). **D** Box plots show the abundance of the significantly different cell populations from the edgeR analysis of the individual samples from HD and CVD patients. Data were analyzed using Mann-Whitney non-parametric test. Significant differences between the two patient cohorts are marked with stars (* > 0.05; ** > 0.01; *** > 0.001, **** > 0.0001)

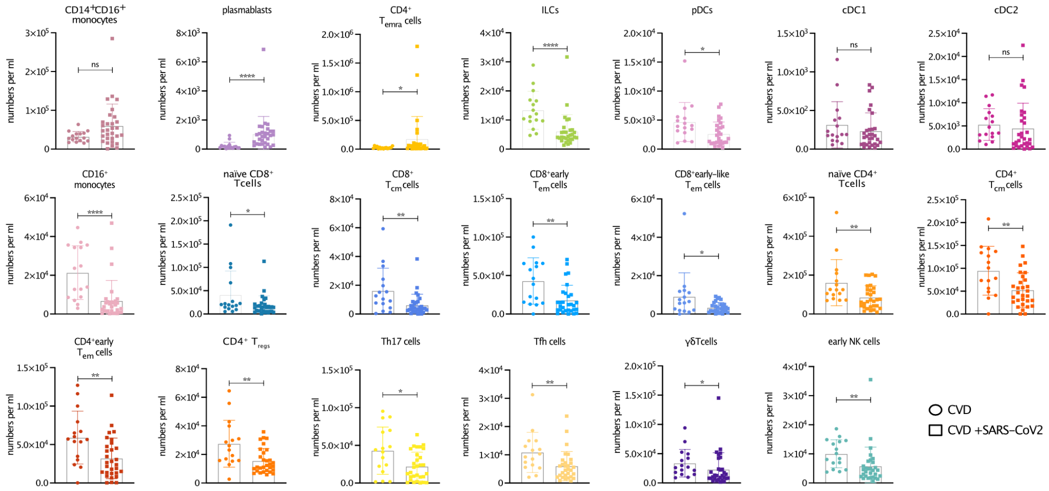


**Supplemental Figure 6: Differences between immune cell populations in CVD patients and SARS-CoV-2-infected CVD patients**
Box plots show the abundance of the depicted cell populations defined by manual gating (Figure S2) of the individual samples from CVD and SARS-CoV-2-infected CVD patients. Data were analyzed using Mann-Whitney non-parametric test. Significant differences between the two patient cohorts are marked with stars (* > 0.05; ** > 0.01; *** > 0.001; **** > 0.0001).

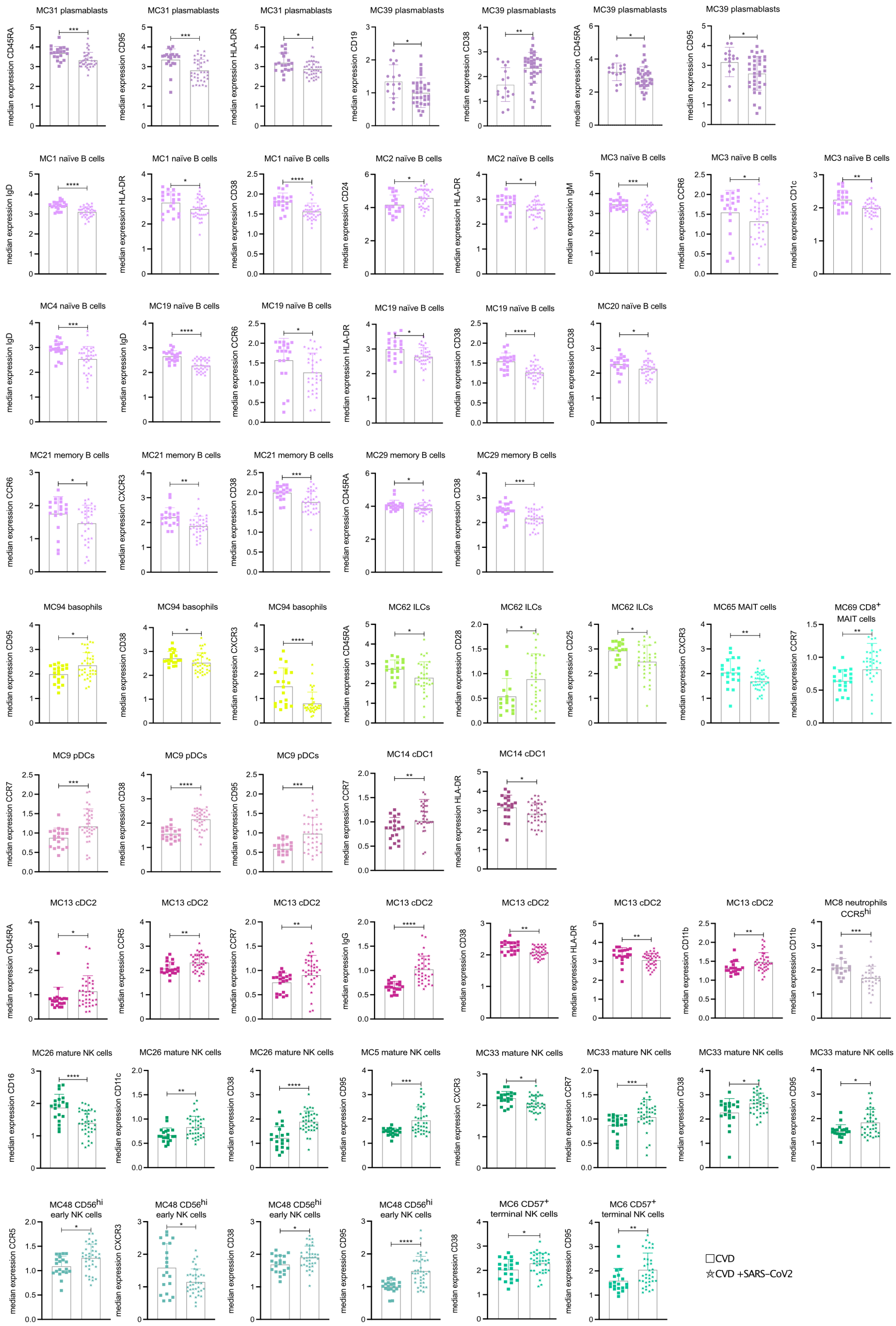


**Supplemental Figure 7: Different marker expression on peripheral immune cell populations from CVD patients and SARS-CoV-2-infected CVD patients**

Boxplots show depicted marker expression on various immune cell MCs (see Figure 2) from CVD (16) and SARS-CoV-2-infected CVD (31) samples. Data were analyzed using Mann-Whitney non-parametric test. Significant differences between the two patient cohorts are marked with stars (* > 0.05; ** > 0.01; *** > 0.001; **** > 0.0001). To be continued on the next two pages.

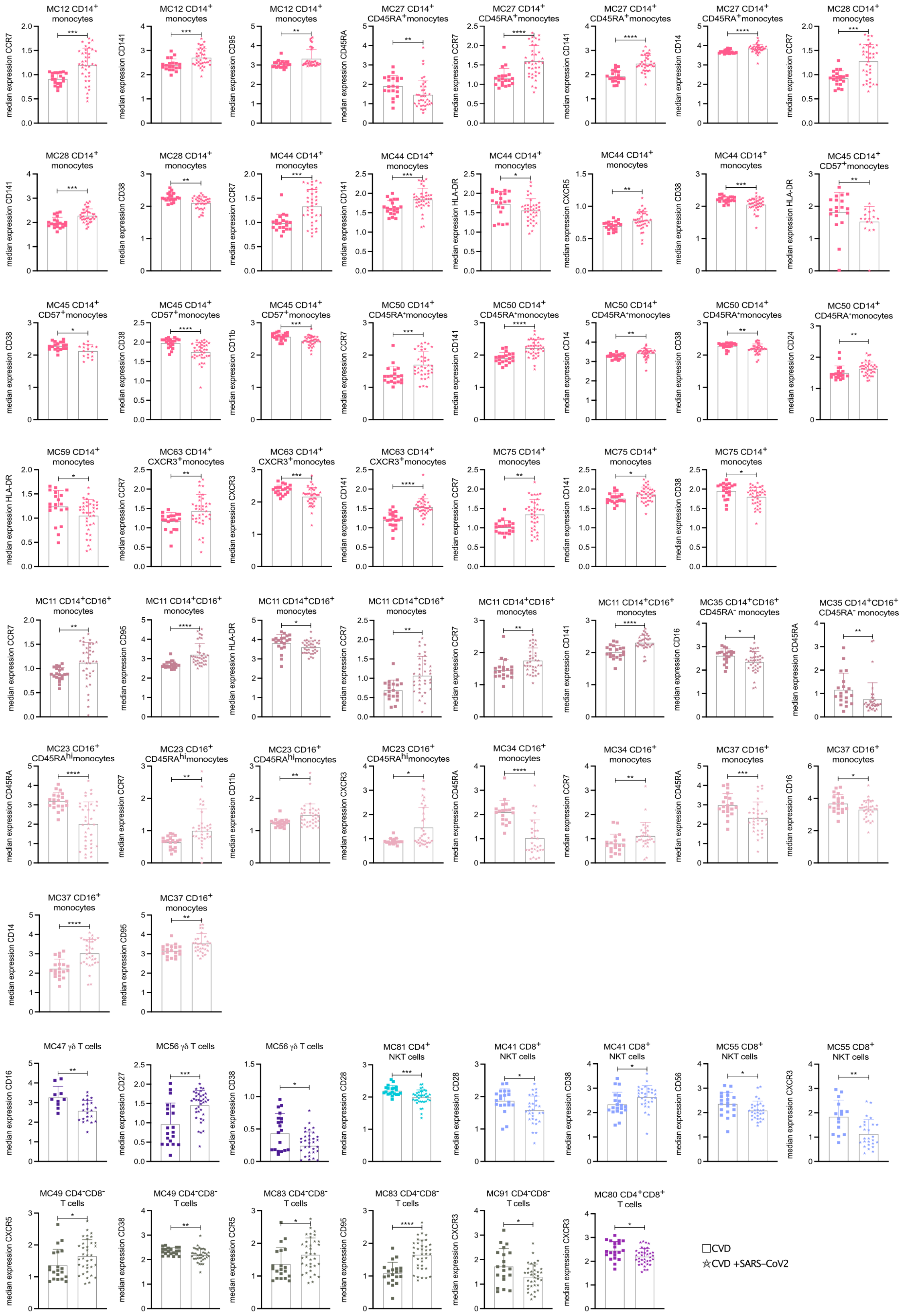

MC12 CD14+ monocytes
median expression CCR7
MC12 CD14+ monocytes
median expression CD141
MC12 CD14+ monocytes
median expression CD95
MC27 CD14+ CD45RA+monocytes
median expression CD45RA
MC27 CD14+ CD45RA+monocytes
median expression CCR7
MC27 CD14+ CD45RA+monocytes
median expression CD141
MC27 CD14+ CD45RA+monocytes
median expression CD14
MC28 CD14+ monocytes
median expression CCR7
MC28 CD14+ monocytes
median expression CD141
MC28 CD14+ monocytes
median expression CD38
MC44 CD14+ monocytes
median expression CCR7
MC44 CD14+ monocytes
median expression CD141
MC44 CD14+ monocytes
median expression HLA-DR
MC44 CD14+ monocytes
median expression CXCR5
MC44 CD14+ monocytes
median expression CD38
MC45 CD14+ CD57+monocytes
median expression HLA-DR
MC45 CD14+ CD57+monocytes
median expression CD38
MC45 CD14+ CD57+monocytes
median expression CD38
MC45 CD14+ CD57+monocytes
median expression CD11b
MC50 CD14+ CD45RA-monocytes
median expression CCR7
MC50 CD14+ CD45RA-monocytes
median expression CD141
MC50 CD14+ CD45RA-monocytes
median expression CD14
MC50 CD14+ CD45RA-monocytes
median expression CD38
MC50 CD14+ CD45RA-monocytes
median expression CD24
MC59 CD14+ monocytes
median expression HLA-DR
MC63 CD14+ CXCR3+monocytes
median expression CCR7
MC63 CD14+ CXCR3+monocytes
median expression CXCR3
MC63 CD14+ CXCR3+monocytes
median expression CD141
MC75 CD14+ monocytes
median expression CCR7
MC75 CD14+ monocytes
median expression CD141
MC75 CD14+ monocytes
median expression CD38
MC11 CD14+CD16+ monocytes
median expression CCR7
MC11 CD14+CD16+ monocytes
median expression CD95
MC11 CD14+CD16+ monocytes
median expression HLA-DR
MC11 CD14+CD16+ monocytes
median expression CCR7
MC11 CD14+CD16+ monocytes
median expression CCR7
MC11 CD14+CD16+ monocytes
median expression CD141
MC35 CD14+CD16+ CD45RA- monocytes
median expression CD16
MC35 CD14+CD16+ CD45RA- monocytes
median expression CD45RA
MC23 CD16+ CD45RAhimonocytes
median expression CD45RA
MC23 CD16+ CD45RAhimonocytes
median expression CCR7
MC23 CD16+ CD45RAhimonocytes
median expression CD11b
MC23 CD16+ CD45RAhimonocytes
median expression CXCR3
MC34 CD16+ monocytes
median expression CD45RA
MC34 CD16+ monocytes
median expression CCR7
MC37 CD16+ monocytes
median expression CD45RA
MC37 CD16+ monocytes
median expression CD16
MC37 CD16+ monocytes
median expression CD14
MC37 CD16+ monocytes
median expression CD95
MC47 γδ T cells
median expression CD16
MC56 γδ T cells
median expression CD27
MC56 γδ T cells
median expression CD38
MC81 CD4+ NKT cells
median expression CD28
MC41 CD8+ NKT cells
median expression CD28
MC41 CD8+ NKT cells
median expression CD38
MC55 CD8+ NKT cells
median expression CD56
MC55 CD8+ NKT cells
median expression CXCR3
MC49 CD4-CD8- T cells
median expression CXCR5
MC49 CD4-CD8- T cells
median expression CD38
MC83 CD4-CD8- T cells
median expression CCR5
MC83 CD4-CD8- T cells
median expression CD95
MC91 CD4-CD8- T cells
median expression CXCR3
MC80 CD4+CD8+ T cells
median expression CXCR3
CVD
CVD +SARS-CoV2

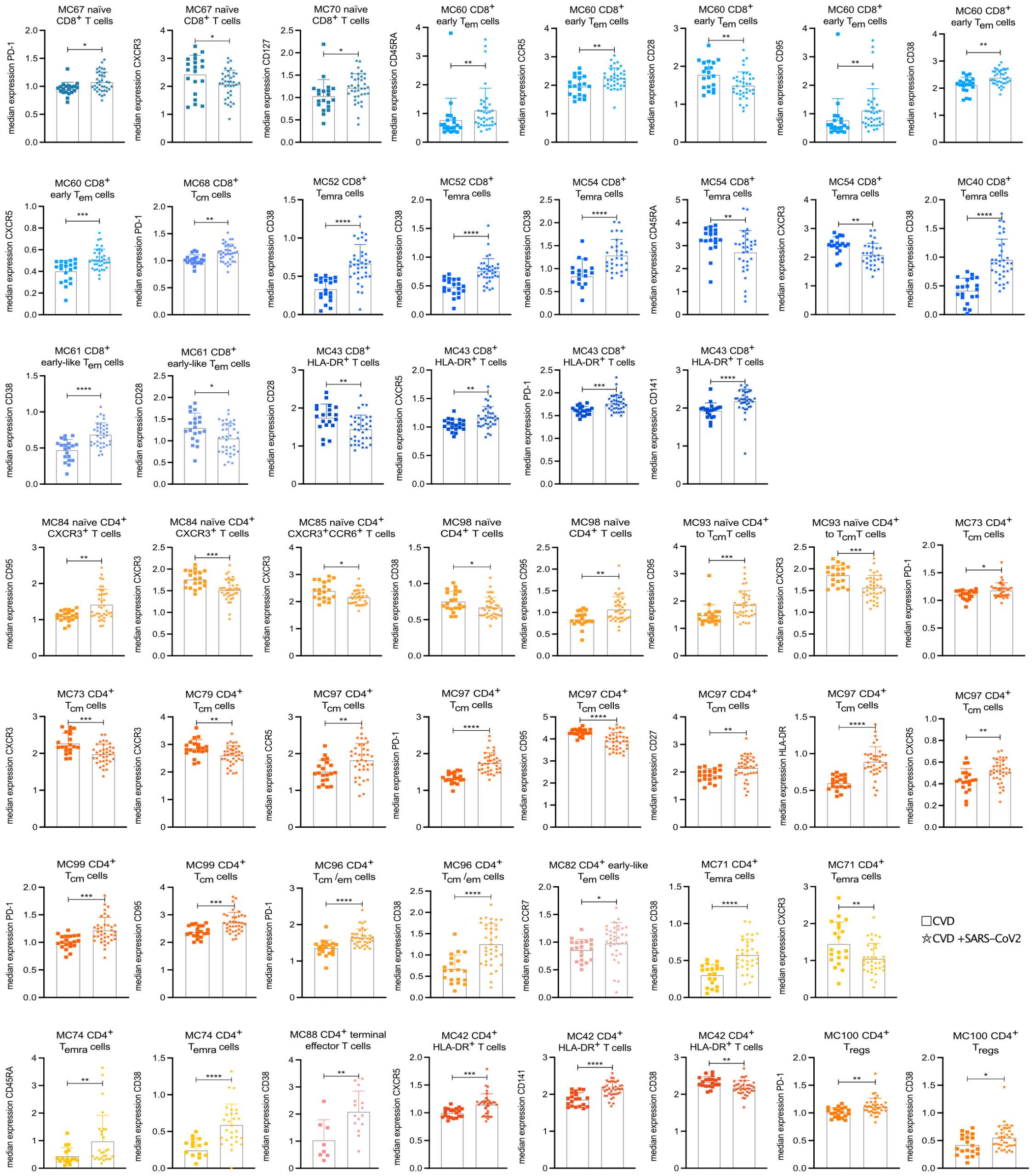

MC67 naïve CD8+ T cells
median expression PD-1
MC67 naïve CD8+ T cells
median expression CXCR3
MC70 naïve CD8+ T cells
median expression CD127
MC60 CD8+ early Tem cells
median expression CD45RA
MC60 CD8+ early Tem cells
median expression CCR5
MC60 CD8+ early Tem cells
median expression CD28
MC60 CD8+ early Tem cells
median expression CD95
MC60 CD8+ early Tem cells
median expression CD38
MC60 CD8+ early Tem cells
median expression CXCR5
MC68 CD8+ Tcm cells
median expression PD-1
MC52 CD8+ Temra cells
median expression CD38
MC52 CD8+ Temra cells
median expression CD38
MC54 CD8+ Temra cells
median expression CD38
MC54 CD8+ Temra cells
median expression CD45RA
MC54 CD8+ Temra cells
median expression CXCR3
MC40 CD8+ Temra cells
median expression CD38
MC61 CD8+ early-like Tem cells
median expression CD38
MC61 CD8+ early-like Tem cells
median expression CD28
MC43 CD8+ HLA-DR+ T cells
median expression CD28
MC43 CD8+ HLA-DR+ T cells
median expression CXCR5
MC43 CD8+ HLA-DR+ T cells
median expression PD-1
MC43 CD8+ HLA-DR+ T cells
median expression CD141
MC84 naïve CD4+ CXCR3+ T cells
median expression CD95
MC84 naïve CD4+ CXCR3+ T cells
median expression CXCR3
MC85 naïve CD4+ CXCR3+CCR6+ T cells
median expression CXCR3
MC98 naïve CD4+ T cells
median expression CD38
MC98 naïve CD4+ T cells
median expression CD95
MC93 naïve CD4+ to TcmT cells
median expression CD95
MC93 naïve CD4+ to TcmT cells
median expression CXCR3
MC73 CD4+ Tcm cells
median expression PD-1
MC73 CD4+ Tcm cells
median expression CXCR3
MC79 CD4+ Tcm cells
median expression CXCR3
MC97 CD4+ Tcm cells
median expression CCR5
MC97 CD4+ Tcm cells
median expression PD-1
MC97 CD4+ Tcm cells
median expression CD95
MC97 CD4+ Tcm cells
median expression CD27
MC97 CD4+ Tcm cells
median expression HLA-DR
MC97 CD4+ Tcm cells
median expression CXCR5
MC99 CD4+ Tcm cells
median expression PD-1
MC99 CD4+ Tcm cells
median expression CD95
MC96 CD4+ Tcm /em cells
median expression PD-1
MC96 CD4+ Tcm /em cells
median expression CD38
MC82 CD4+ early-like Tem cells
median expression CCR7
MC71 CD4+ Temra cells
median expression CD38
MC71 CD4+ Temra cells
median expression CXCR3
CVD
CVD +SARS-CoV2
MC74 CD4+ Temra cells
median expression CD45RA
MC74 CD4+ Temra cells
median expression CD38
MC88 CD4+ terminal effector T cells
median expression CD38
MC42 CD4+ HLA-DR+ T cells
median expression CXCR5
MC42 CD4+ HLA-DR+ T cells
median expression CD141
MC42 CD4+ HLA-DR+ T cells
median expression CD38
MC100 CD4+ Tregs
median expression PD-1
MC100 CD4+ Tregs
median expression CD38

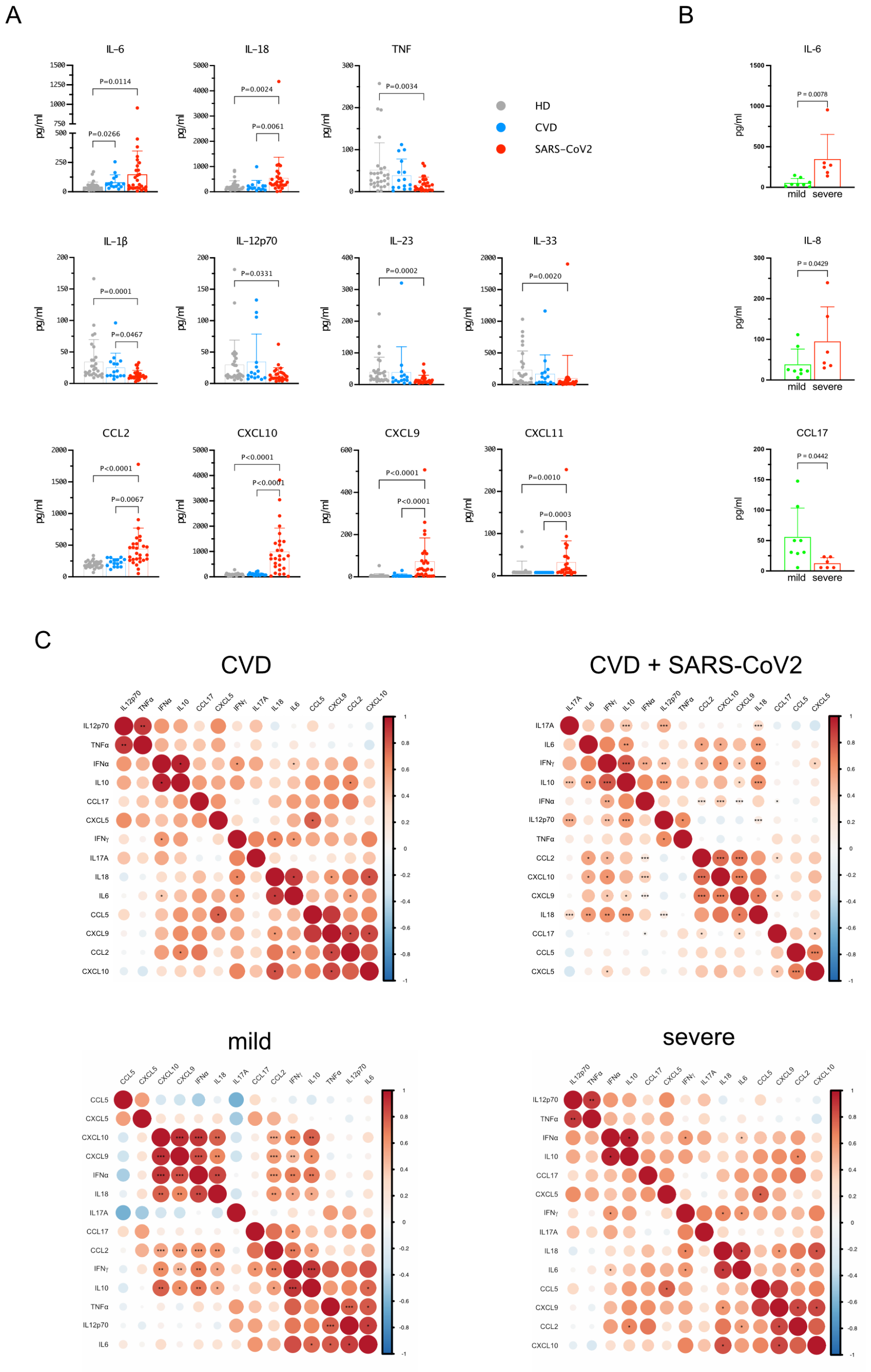


**Supplemental Figure 8: Cytokine expression in the plasma**
Box plots show depicted cytokine levels in the plasma from HD (28), CVD patients (15) and SARS-CoV-2-infected CVD patients (28) (**A**) or mild (10) and severe (7) SARS-CoV-2-infected CVD patients (**B**). (**C**) Clustered correlation analysis of the plasma cytokines/chemokines levels in the depicted patient cohorts. The size of the circle reflects the correlation coefficient either negative (blue) or positive (red). Data were analyzed using Kruskal-Wallis non-parametric test with Dunn's post-test (**A**) or nonparametric Mann-Whitney test (**B**) or Spearman correlation coefficient (**C**). Significant P values are depicted above the lines (* > 0.05; ** > 0.01; *** > 0.001; **** > 0.0001).

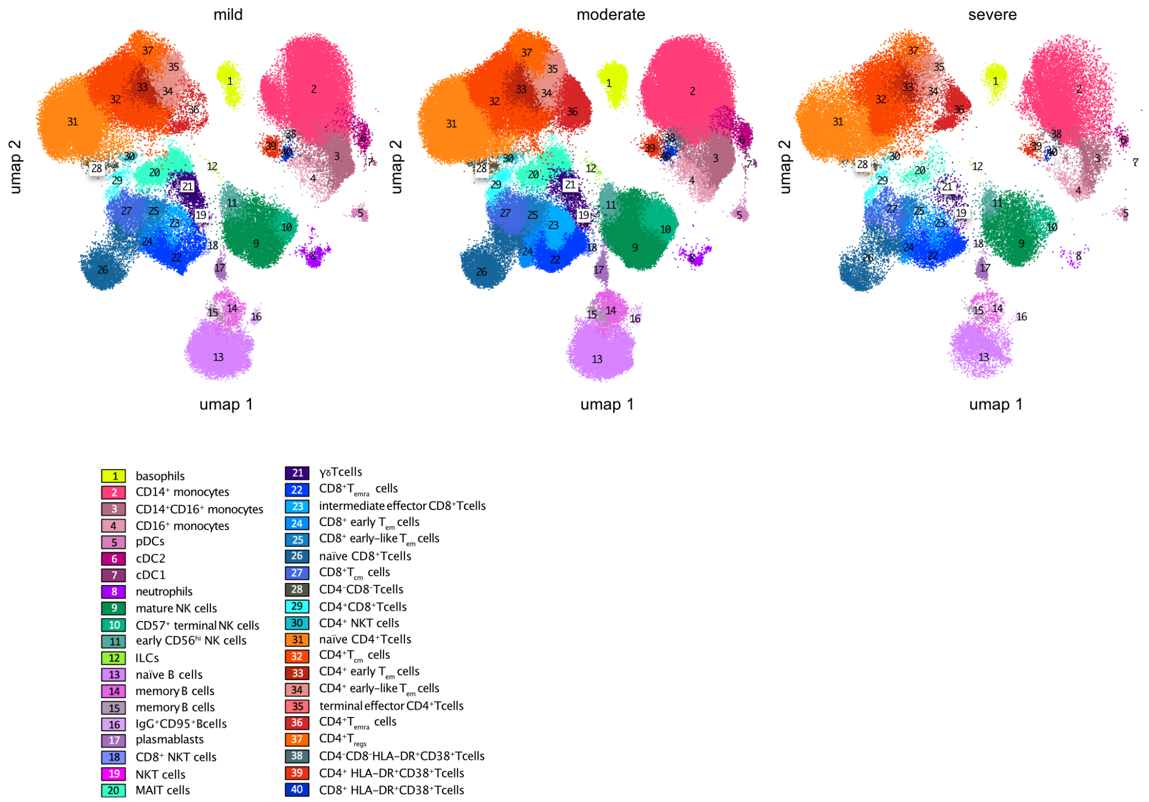


**Supplemental Figure 9:**
High-dimensional data analysis of PBMCs displaying 40 FlowSOM clusters projected onto two UMAP dimensions. The UMAP plots show concatenated events from PBMC samples from CVD + SARS-CoV-2-infected patients (37) regarding severity (mild (10), moderate (20), and severe (7)).

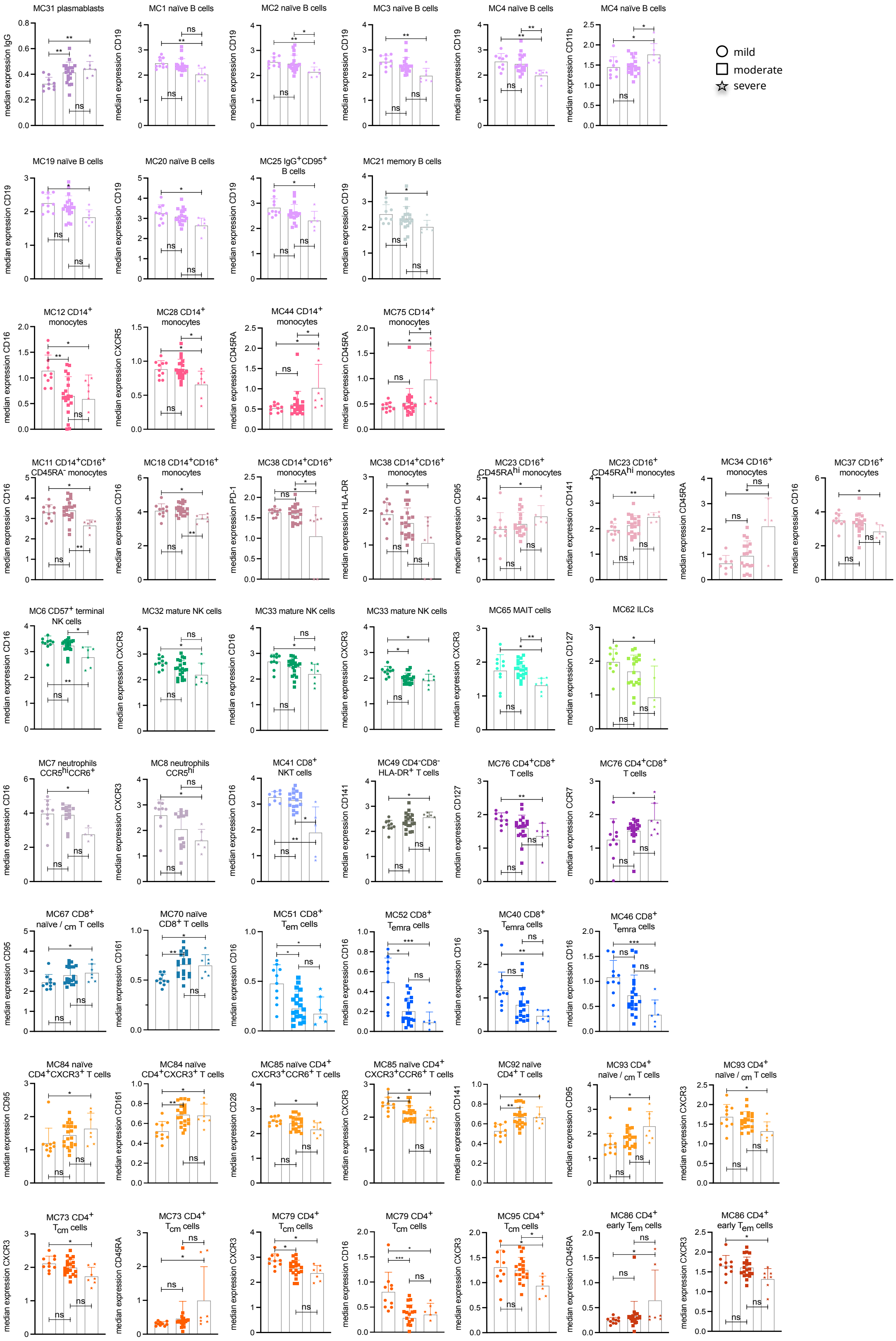


**Supplemental Figure 10: Different marker expression on immune cell populations in mild, moderate, and severe SARS-CoV-2-infected CVD patients**

Boxplots show depicted marker expression on various immune cell MCs (see Figure 2) from mild (10), moderate (20), and severe (7) SARS-CoV-2-infected CVD patients. Data were analyzed using Kruskal-Wallis non-parametric test with Dunn's post-test. Significant differences between the two patient cohorts are marked with stars (* > 0.05; ** > 0.01; *** > 0.001).

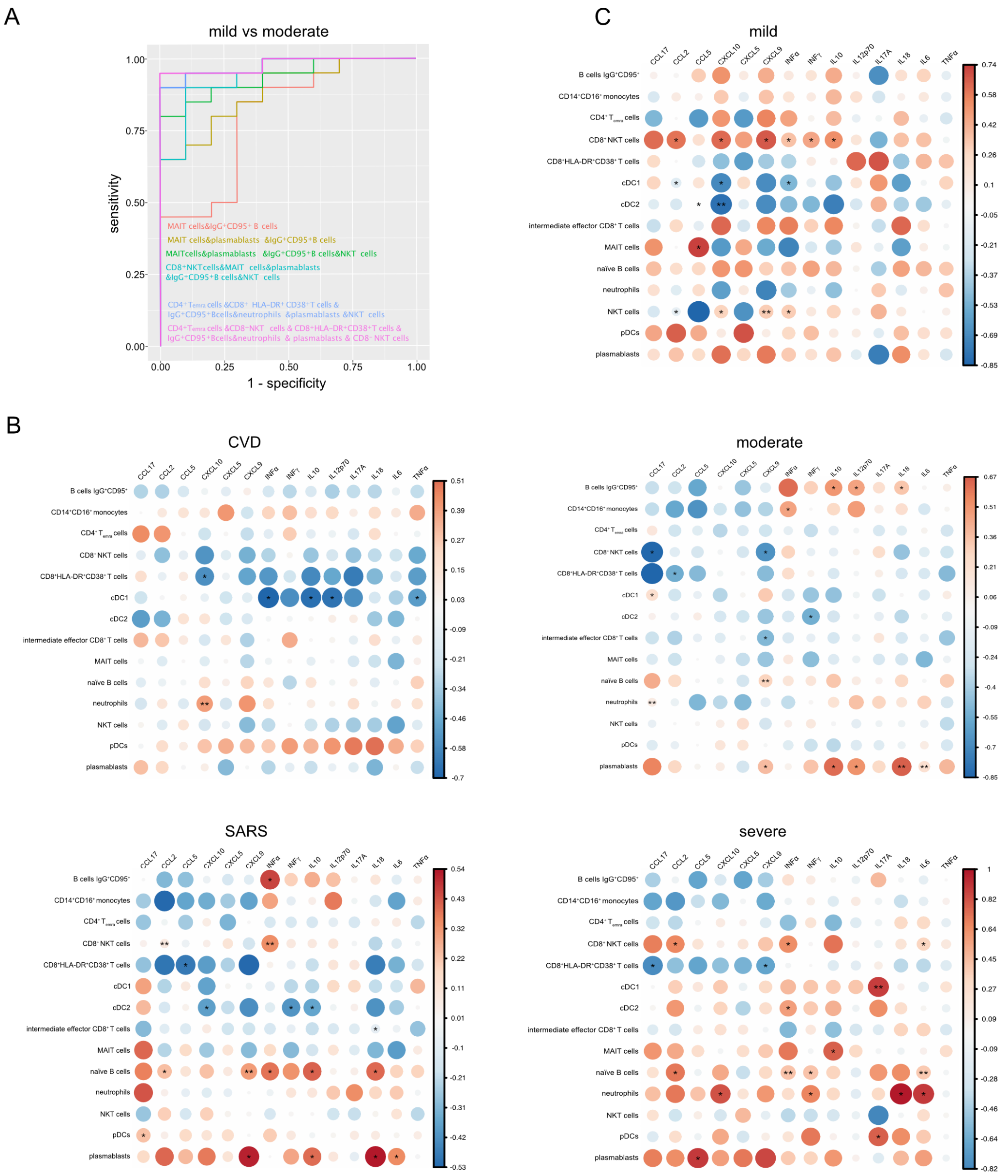


**Supplemental Figure 11: Combined analysis of cell populations and cytokines/chemokines**
(A) CombiROC analysis of cell populations in mild vs moderately SARS-CoV-2-infected CVD patients. Correlation analysis of plasma cytokines/chemokines levels and the depicted cell populations in CVD patients and SARS-CoV-2-infected CVD patients (B) or mild, moderate, and severely SARS-CoV-2-infected CVD patients (C). The size of the circle reflects the correlation coefficient either negative (blue) or positive (red). Data were analyzed using nonparametric Mann-Whitney test (A) or Spearman correlation coefficient (B). Significant P values are depicted (* > 0.05; ** > 0.01; *** > 0.001; **** > 0.0001).

**Table S1. Baseline characteristics of CVD patients at study entry stratified by presence of acute SARS-CoV-2 infection**

| Parameters | All patients (n=57) | CVD (n=20) | CVD-SARS-CoV-2 (n=37) | p-Value |
|---|---|---|---|---|
| Clinical characteristics | | | | |
| Age (y) | 68 (58-80) | 67 (59-78) | 69 (57-81) | 0.647 |
| Male | 29 (50.9) | 9 (45) | 20 (54.1) | 0.514 |
| BMI (kg/m²) | 26.6 (24.9-29.6) | 25 (23.3-27.3) | 27.4 (25.1-31.4) | 0.091 |
| ICU admission | 11 (19.3) | 0 (0) | 11 (29.7) | **0.007** |
| ARDS | | | | |
| - mild | 19 (33.3) | 0 (0) | 19 (51.4) | |
| - moderate | 8 (14) | 0 (0) | 8 (21.6) | **<0.001** |
| - severe | 5 (8.7) | 0 (0) | 5 (13.5) | |
| Horovitz Index | | | | |
| - HI > 300 mmHg | 33 (57.9) | 20 (100) | 13 (35.1) | |
| - HI 201 - 300 mmHg | 4 (7) | 0 (0) | 4 (10.8) | **<0.001** |
| - HI 101 - 200 mmHg | 11 (19.3) | 0 (0) | 11 (29.7) | |
| - HI ≤ 100 mmHg | 9 (15.8) | 0 (0) | 9 (24.3) | |
| High flow $0_2$ Therapy | 5 (8.1) | 0 (0) | 5 (13.5) | |
| Mechanical ventilation | 8 (14) | 0 (0) | 8 (21.6) | |
| Vasopressor | 6 (10.5) | 0 (0) | 6 (16.2) | |
| Lymphocyte count at Nadir (1000/µl) | 0.8 (0.6-1.2) | | 0.8 (0.6-1.2) | |
| Bacterial Co-Infection | 14 (24.6) | 0 (0) | 14 (37.8) | **0.025** |
| Dialysis | 5 (8.1) | 0 (0) | 5 (13.5) | |
| Acute hepatic injury | 4 (7) | 0 (0) | 4 (10.8) | |
| Symptoms on admission | | | | |
| Cough | 18 (31.6) | 0 (0) | 18 (48.6) | **<0.001** |
| Dyspnea | 18 (31.6) | 0 (0) | 18 (48.6) | **<0.001** |
| Fever | 20 (35.1) | 0 (0) | 20 (54.1) | **<0.001** |
| Cardiovascular risk factors | | | | |
| Arterial hypertension | 42 (73.7) | 15 (75) | 27 (73) | 0.868 |
| Dyslipidemia | 29 (50.9) | 9 (45) | 20 (54.1) | 0.449 |
| Diabetes mellitus | 10 (17.5) | 2 (10) | 8 (14.) | 0.271 |

| | | | | |
|---|---|---|---|---|
| Current smokers | 7 (12.3) | 3 (15) | 4 (10.8) | 0.673 |
| Obesity | 18 (31.6) | 5 (25) | 13 (35.1) | 0.393 |
| Atrial fibrillation | 14 (24.6) | 6 (30) | 8 (13.5) | 0.520 |
| Chronic kidney disease | 7 (12.3) | 1 (5) | 6 (16.2) | 0.206 |
| COPD | 3 (5.3) | 0 (0) | 3 (8.1) | 0.184 |
| Malignoma | 8 (14.1) | 5 (25) | 3 (8.1) | 0.088 |
| NYHA | | | | |
| - 1 | 10 (17.5) | 8 (40) | 2 (5.4) | |
| - 2 | 2 (3.5) | 2 (10) | 0 (0) | **0.001** |
| - 3 | 2 (3.5) | 1 (5) | 1 (2.7) | |
| Parameters of echocardiography | | | | |
| Left ventricular hypertrophy | 22 (38.6) | 5 (25) | 17 (45.9) | 0.062 |
| Right ventricular function | 3 (5.3) | 0 (0) | 3 (8.1) | 0.433 |
| Right ventricular dilatation | 13 (22.8) | 1 (5) | 12 (32.4) | 0.057 |
| PE | 17 (29.8) | 0 (0) | 17 (45.9) | **0.003** |
| Pleural effusion | 10 (17.6) | 1 (5) | 9 (24.3) | 0.676 |
| Bilateral nodular opacities | 19 (33.3) | 0 (0) | 19 (51.4) | 0.328 |
| Ground-glass opacities | 9 (15.8) | 0 (0) | 9 (24.3) | 0.410 |
| Peribronchial thickening | 2 (3.5) | 0 (0) | 2 (5.4) | 0.732 |
| Focal consolidations | 9 (15.8) | 0 (0) | 9 (24.3) | 0.410 |
| Venous congestion | 6 (10.5) | 0 (0) | 6 (16.2) | 0.522 |
| Atelectasis | 1 (1.8) | 0 (0) | 1 (2.7) | 0.809 |
| Parameters of electrocardiography | | | | |
| Heart Rate (bpm) | 76 (67-84) | 69 (64.3-82.8) | 77 (67.5-85.5) | 0.311 |
| Systolic blood pressure (mmHg) | 140 (130-151.3) | 145 (130-165) | 140 (125-150) | 0.372 |
| Heart Rhythm | | | | |
| - Sinus rhythm | 48 (90.4) | 18 (90.4) | 30 (90.4) | |
| - Atrial fibrillation | 4 (7) | 1 (5) | 3 (8.1) | 0.468 |
| - PM | 1 (1.8) | 0 (0) | 1 (2.7) | |
| - SVT | 1 (1.8) | 1 (5) | 0 (0) | |
| Laboratory parameters and biomarkers | | | | |
| Leukocytes (1000/µL) | 6945 (4900-8925) | 7415 (6010-8747.5) | 6265 (4215-8965) | 0.700 |

| | | | | |
|---|---|---|---|---|
| Lymphocytes (1000/µL) | 1000 (665-1530) | 1540 (1235-2135) | 845 (610-1257) | **<0.001** |
| Hb (g/dL) | 13.1 (12.0-13.9) | 13.5 (12.6-13.9) | 13.1 (11.9-13.8) | 0.449 |
| Platelets (1000/µL) | 211.5 (160-297.8) | 262.5 (203.5-298.8) | 176.5 (147-284.3) | 0.094 |
| INR (%) | 1 (1-1.1) | 1 (1-1.1) | 1 (1-1.2) | 0.289 |
| PTT (s) | 24 (22-27) | 24 (23-27) | 24 (22-27) | 0.514 |
| D-Dimer (µg/dL) | 0.8 (0.6-1.8) | 0.31 (0.31-0.31) | 0.9 (0.6-1.8) | 0.310 |
| Creatinine (mg/dL) | 0.8 (0.7-1.1) | 0.8 (0.6-0.9) | 0.9 (0.7-1.3) | 0.100 |
| GFR-MDRD (ml/m$^2$) | 79.8 (52.8-101.8) | | 79.8 (52.8-101.8) | |
| Sodium (mmol/L) | 139 (136-140) | 140 (139-141) | 137 (134.5-139) | **<0.001** |
| CRP (mg/dL) | 1.4 (0.3-6.3) | 0.2 (0.03-1) | 3.1 (1.3-11.2) | **<0.001** |
| PCT (ng/mL) | 0.1 (0.1-0.2) | | 0.1 (0.1-0.2) | |
| IL-6 | 24.3 (8.7-34.5) | | 24.3 (8.7-34.5) | |
| hs TNI (ng/dL) | 8 (3.8-18.5) | 5 (3.3-7.8) | 10.5 (5.3-22) | **0.026** |
| NT-pro-BNP (ng/L) | 410 (121-1349.3) | | 519 (128.8-1487.5) | 0.144 |
| CK (U/L) | 110.5 (65.3-185.5) | 84.5 (55-132) | 121.5 (66.3-203.3) | 0.094 |
| AST (U/L) | 32 (17-46) | 17 (12.5-21) | 41 (21-54) | **<0.001** |
| ALT (U/L) | 24 (16.3-39) | 19 (14.3-32) | 30.5 (19-41.8) | **0.042** |
| LDH (U/L) | 225 (191-290) | 195 (172.3-209.3) | 264 (218-361) | **<0.001** |
| HbA1c (%) | 6.1 (5.8-6.5) | 5.9 (5.5-6.0) | 6.3 (5.8-6.5) | **0.011** |
| Concomitant cardiac medication at study entry | | | | |
| Oral anticoagulation | 11 (19.3) | 6 (30) | 5 (13.5) | 0.215 |
| ACE-I or ARB | 36 (63.2) | 13 (65) | 23 (62.2) | 0.563 |
| Diuretics | 15 (26.3) | 3 (15) | 12 (32.4) | 0.147 |
| Calcium channel blockers | 12 (21.1) | 5 (25) | 7 (12.8) | 0.594 |
| Beta blockers | 23 (40.4) | 6 (30) | 17 (18.9) | 0.263 |
| Statins | 29 (50.9) | 10 (50) | 19 (51.4) | 0.992 |
| ASA | 23 (40.1) | 10 (50) | 13 (35.1) | 0.238 |
| P2Y12-Inhibitor | 5 (8.8) | 4 (20) | 1 (2.7) | 0.165 |

**Table S1** Values are n (%) or are given as median and interquartile range (IQR). ACE - Angiotensin Converting Enzyme. Afib – atrial fibrillation. ALT – alanine amino-tranferase. ARB – Angiotensin II receptor blockers. ARDS – Acute respiratory distress syndrome. ASA – Acetylsalicylic acid. AST – aspartate-aminotransferase. BMI – body mass index. CAD – coronary artery disease. CK – creatinine kinase. COPD – chronic obstructive pulmonary disease. CVD – cardio vascular disease. CRP – C-reactive protein. GFR-MDRD – glomerular filtration rate. Hb – hemoglobin. HI – Horovitz Index. hs TNI – high sensitive Troponin I. ICU – Intensive care unit. INR – international normalized ratio. LDH – lactate dehydrogenase. NT-pro-BNP – N-terminal pro- brain natriuretic peptide. NYHA – New York Heart Association. PCT – procalcitonin. PE – pericardial effusion. PM – Pacemaker. PTT – partial thromboplastin time. SARS-CoV-2 – severe acute respiratory syndrome coronavirus-2. SVT – supraventricular tachycardia.

**Table S2. Baseline characteristics of SARS-CoV-2 infected CVD patients at study entry stratified by ISARIC-WHO-4C-Mortality-Score**

| Parameters | All patients (n=37) | Mild (n=10) | Moderate (n=20) | Severe (n=7) | p-Value |
|---|---|---|---|---|---|
| Clinical characteristics | | | | | |
| Age (y) | 69 (57-81) | 60 (53-68) | 79 (58-81) | 70 (64-84) | **0.016** |
| Male | 20 (54.1) | 6 (60) | 9 (45) | 5 (71.4) | 0.438 |
| BMI (kg/m²) | 27.4 (25.1-31.4) | 29.4 (27.7-34.8) | 26.2 (24.4-31) | 25.6 (25-28.7) | **0.022** |
| ICU admission | 11 (29.7) | 0 (0) | 4 (20) | 7 (100) | **<0.001** |
| ARDS | | | | | |
| - mild | 19 (51.4) | 7 (70) | 11 (55) | 1 (14.3) | |
| - moderate | 8 (21.6) | 1 (10) | 5 (25) | 2 (28.6) | **0.010** |
| - severe | 5 (13.5) | 0 (0) | 1 (5) | 4 (57.1) | |
| Horovitz Index | | | | | |
| - HI > 300 mmHg | 13 (35.1) | 10 (100) | 3 (15) | 0 (0) | |
| - HI 201 - 300 mmHg | 4 (10.8) | 0 (0) | 4 (20) | 0 (0) | **<0.001** |
| - HI 101 - 200 mmHg | 11 (29.7) | 0 (0) | 11 (55) | 0 (0) | |
| - HI ≤ 100 mmHg | 9 (24.3) | 0 (0) | 2 (10) | 7 (100) | |
| High flow $0_2$ Therapy | 5 (13.5) | 0 (0) | 1 (5) | 4 (57.1) | **0.002** |
| Mechanical ventilation | 8 (21.6) | 0 (0) | 3 (15) | 5 (71.4) | **0.002** |
| Vasopressor | 6 (16.2) | 0 (0) | 2 (10) | 4 (57.1) | **0.009** |
| Lymphocyte count at Nadir (1000/µl) | 0.8 (0.6-1.2) | 0.9 (0.6-1.3) | 0.8 (0.6-1.6) | 0.6 (0.5-1) | 0.505 |
| Bacterial Co-Infection | 14 (37.8) | 2 (20) | 6 (30) | 6 (85.7) | **0.012** |
| Dialysis | 5 (13.5) | 1 (10) | 3 (15) | 1 (14.3) | 0.255 |
| Acute hepatic injury | 4 (10.8) | 0 (0) | 3 (15) | 1 (14.3) | 0.451 |
| Symptoms on admission | | | | | |
| Cough | 18 (48.6) | 3 (30) | 11 (55) | 4 (57.1) | 0.330 |
| Dyspnea | 18 (48.6) | 5 (50) | 10 (50) | 3 (42.9) | 0.907 |
| Fever | 20 (54.1) | 6 (60) | 9 (45) | 5 (71.4) | 0.519 |
| Cardiovascular risk factors | | | | | |
| Arterial hypertension | 27 (73) | 7 (70) | 14 (70) | 6 (85.7) | 0.701 |
| Dyslipidemia | 20 (54.1) | 7 (70) | 11 (55) | 2 (28.6) | 0.229 |

| | | | | | |
|---|---|---|---|---|---|
| Diabetes mellitus | 8 (14.) | 2 (20) | 4 (20) | 2 (28.6) | 0.884 |
| Current smokers | 4 (10.8) | 2 (20) | 1 (5) | 1 (14.3) | 0.465 |
| Obesity | 13 (35.1) | 5 (50) | 6 (30) | 2 (28.6) | 0.555 |
| Atrial fibrillation | 8 (13.5) | 1 (10) | 4 (20) | 3 (42.9) | 0.272 |
| COPD | 3 (8.1) | 0 (0) | 2 (10) | 1 (14.3) | 0.508 |
| Malignoma | 3 (8.1) | 0 (0) | 2 (10) | 1 (14.3) | 0.508 |
| NYHA | | | | | |
| - 1 | 2 (5.4) | 0 (0) | 1 (5) | 1 (14.3) | |
| - 2 | 0 (0) | 0 (0) | 0 (0) | 0 (0) | 0.640 |
| - 3 | 1 (2.7) | 0 (0) | 1 (5) | 0 (0) | |
| Parameters of echocardiography | | | | | |
| Right ventricular function | 3 (8.1) | 0 (0) | 2 (10) | 1 (14.3) | 0.640 |
| Right ventricular dilatation | 12 (32.4) | 4 (40) | 7 (35) | 1 (14.3) | 0.729 |
| PE | 17 (45.9) | 1 (10) | 12 (60) | 4 (57.1) | 0.097 |
| Pleural effusion | 9 (24.3) | 2 (20) | 4 (20) | 3 (42.9) | 0.345 |
| Bilateral nodular opacities | 19 (51.4) | 6 (60) | 9 (45) | 4 (57.1) | 0.472 |
| Ground-glass opacities | 9 (24.3) | 2 (20) | 4 (20) | 3 (42.9) | 0.509 |
| Peribronchial thickening | 2 (5.4) | 1 (10) | 0 (0) | 1 (14.3) | 0.256 |
| Focal consolidations | 9 (24.3) | 3 (30) | 4 (20) | 2 (28.6) | 0.771 |
| Venous congestion | 6 (16.2) | 0 (0) | 4 (20) | 2 (28.6) | 0.258 |
| Atelectasis | 1 (2.7) | 0 (0) | 0 (0) | 1 (14.3) | 0.128 |
| Parameters of electrocardiography | | | | | |
| Heart Rate (bpm) | 77 (67.5-85.5) | 73 (66.3-85.5) | 77.5 (68.3-84.3) | 79 (72-92) | 0.773 |
| Systolic blood pressure (mmHg) | 140 (125-150) | 137.5 (120-142.5) | 145 (119.3-151.3) | 140 (130-147) | 0.145 |
| Heart Rhythm | | | | | |
| - Sinus rhythm | 30 (90.4) | 8 (80) | 15 (75) | 7 (100) | |
| - Atrial fibrillation | 3 (8.1) | 1 (10) | 2 (10) | 0 (0) | 0.768 |
| - PM | 1 (2.7) | 0 (0) | 1 (5) | 0 (0) | |
| - SVT | 0 (0) | 0 (0) | 0 (0) | 0 (0) | |
| Laboratory parameters and biomarkers | | | | | |

| | | | | | |
|---|---|---|---|---|---|
| Leukocytes (1000/µL) | 6265 (4215-8965) | 6935 (3552.5-10635) | 5940 (4590-7740) | 6970 (4290-9810) | 0.907 |
| Lymphocytes (1000/µL) | 845 (610-1257) | 1095 (735-1360) | 890 (670-1520) | 530 (410-650) | **0.011** |
| Hb (g/dL) | 13.1 (11.9-13.8) | 13.6 (11.3-14.4) | 13 (12-13.4) | 13.3 (11.9-14) | 0.258 |
| Platelets (1000/µL) | 176.5 (147-284.3) | 192.5 (147-326.8) | 191 (143-246) | 164 (143-334) | 0.907 |
| INR (%) | 1 (1-1.2) | 1.1 (1-1.2) | 1 (1-1.2) | 1 (1-1.1) | 0.636 |
| PTT (s) | 24 (22-27) | 23.5 (22-28) | 23 (22-25) | 26 (23-28) | 0.333 |
| D-Dimer (µg/dL) | 0.9 (0.6-1.8) | 0.31 (0.31-0.31) | 0.9 (0.6-1.8) | 1.7 (0.9-2.2) | 0.576 |
| Creatinine (mg/dL) | 0.9 (0.7-1.3) | 0.9 (0.8-1.4) | 0.9 (0.7-1.1) | 1 (0.6-1.4) | 0.598 |
| GFR-MDRD (ml/m$^2$) | 79.8 (52.8-101.8) | 74.1 (49.6-102.9) | 80.1 (58.5-108.4) | 73.9 (48.3-100.6) | 0.788 |
| Sodium (mmol/L) | 137 (134.5-139) | 138 (136-140) | 136.5 (134.3-139) | 135 (126-140) | 0.699 |
| CRP (mg/dL) | 3.1 (1.3-11.2) | 2.1 (0.6-6.2) | 2.5 (1.3-8.6) | 12.5 (11.9-18.3) | **0.011** |
| PCT (ng/mL) | 0.1 (0.1-0.2) | 0.1 (0-0.2) | 0.1 (0.1-0.2) | 0.2 (0.1-0.4) | 0.128 |
| IL-6 | 24.3 (8.7-34.5) | 9.9 (5.9-18.4) | 21.8 (8.2-36.5) | 34.1 (31.5-63) | **0.007** |
| hs TNI (ng/dL) | 10.5 (5.3-22) | 8 (2.5-38) | 8 (6-22) | 18 (12-26.3) | 0.185 |
| NT-pro-BNP (ng/L) | 519 (128.8-1487.5) | 133 (105.5-626.5) | 595 (119-11911) | 1213 (602-9856.3) | 0.064 |
| CK (U/L) | 121.5 (66.3-203.3) | 91 (62.8-195.3) | 122 (67-217) | 188 (116-207) | 0.230 |
| AST (U/L) | 41 (21-54) | 29.5 (16.3-49) | 39.5 (25.8-55) | 43 (39-62) | 0.774 |
| ALT (U/L) | 30.5 (19-41.8) | 34 (18.5-41.8) | 30 (19-42) | 26 (18-63) | 0.742 |
| LDH (U/L) | 264 (218-361) | 221.5 (174.8-270.5) | 286 (217.3-338.5) | 367 (249-483) | 0.079 |
| HbA1c (%) | 6.3 (5.8-6.5) | 6.1 (5.7-6.6) | 6.1 (5.9-6.4) | 6.4 (6.1-6.9) | 0.621 |
| Concomitant cardiac medication at study entry | | | | | |
| Oral anticoagulation | 5 (13.5) | 2 (20) | 2 (10) | 1 (14.3) | 0.390 |
| ACE-I or ARB | 23 (62.2) | 6 (60) | 12 (60) | 5 (71.4) | 0.087 |
| Diuretics | 12 (32.4) | 4 (40) | 1 (5) | 1 (14.3) | 0.425 |

| | | | | | |
|---|---|---|---|---|---|
| Calcium channel blockers | 7 (12.8) | 2 (20) | 5 (25) | 0 (0) | 0.324 |
| Beta blockers | 17 (18.9) | 5 (50) | 9 (45) | 3 (42.8) | 0.959 |
| Statins | 19 (51.4) | 6 (60) | 10 (50) | 3 (42.8) | 0.784 |
| ASA | 13 (35.1) | 4 (40) | 7 (35) | 2 (28.6) | 0.886 |
| P2Y12-Inhibitor | 1 (2.7) | 0 (0) | 1 (5) | 0 (0) | 0.663 |

**Table S2** Values are n (%) or are given as median and interquartile range (IQR). ACE - Angiotensin Converting Enzyme. Afib – atrial fibrillation. ALT – alanine amino-tranferase. ARB – Angiotensin II receptor blockers. ARDS – Acute respiratory distress syndrome. ASA – Acetylsalicylic acid. AST – aspartate-aminotransferase. BMI – body mass index. CAD – coronary artery disease. CK – creatinine kinase. COPD – chronic obstructive pulmonary disease. CVD – cardio vascular disease. CRP – C-reactive protein. GFR-MDRD – glomerular filtration rate. Hb – hemoglobin. HI – Horovitz Index. hs TNI – high sensitive Troponin I. ICU – Intensive care unit. INR – international normalized ratio. LDH – lactate dehydrogenase. NT-pro-BNP – N-terminal pro- brain natriuretic peptide. NYHA – New York Heart Association. PCT – procalcitonin. PE – pericardial effusion. PM – Pacemaker. PTT – partial thromboplastin time. SARS-CoV-2 – severe acute respiratory syndrome coronavirus-2. SVT – supraventricular tachycardia.

**Table S3 – Antibodies & Materials**

| Antibody | Company | Catalognumber | Clone | Concentration |
|---|---|---|---|---|
| CD1c AlexaFluor647 | BioLegend | 331510 | L161 | 1,25 µg/ml |
| CD3 BV510 | BioLegend | 317332 | OKT3 | 0,75 µg/ml |
| CD4 cFluorYG584 | Cytek Biosciences | R7-20041 | SK3 | 0,25 µg/ml |
| CD8 BUV805 | BD Bioscience | 612889 | SK1 | 0,8 µg/ml |
| CD11b PerCP-Cy5.5 | BioLegend | 301338 | ICRF44 | 5 µg/ml |
| CD11c BUV661 | BD Bioscience | 612967 | B-ly6 | 1,25 µl in 100 µl |
| CD14 Spark Blue 550 | BioLegend | 367148 | 63D3 | 1,6 µg/ml |
| CD16 BUV496 | BD Bioscience | 612944 | 3G8 | 0,6 µg/ml |
| CD19 Spark NIR 685 | BioLegend | 302270 | HIB19 | 0,6 µg/ml |
| CD20 Pacific Orange | Thermo Fisher Scientific | MHCD2030 | HI47 | 1,25 µl in 100 µl |
| CD24 PE Dazzle594 | BioLegend | 311134 | ML5 | 2 µg/ml |
| CD25 PE | BioLegend | 302606 | BC96 | 0,75 µg/ml |
| CD27 APC | BioLegend | 356410 | M-T271 | 2,5 µg/ml |
| CD28 BV650 | BioLegend | 302946 | CD28.2 | 1,5 µg/ml |
| CD38 APC Fire810 | BioLegend | 102746 | HIT2 | 0,75 µg/ml |
| CD45 PerCP | BioLegend | 368506 | 2D1 | 2 µg/ml |
| CD45RA BUV395 | BD Bioscience | 740315 | 5H9 | 2 µg/ml |
| CD56 BUV737 | BD Bioscience | 612766 | NCAM16.2 | 2,5 µl in 100 µl |
| CD57 FITC | BioLegend | 359604 | HNK-1 | 0,6 µg/ml |
| CD95 PE/Cy5 | BioLegend | 305610 | DX2 | 1,2 µg/ml |
| CD123 Super Bright436 | Thermo Fisher Scientific | 62-1239-42 | 6H6 | 0,3 µg/ml |
| CD127 APC-R700 | BD Bioscience | 565185 | HIL-7R-M21 | 1,25 µl in 100 µl |
| CD141 BrilliantBlue515 | BD Bioscience | 566017 | 1A4 | 3 µl from a 1:5 pre-dilution in 100 µl |
| CD161 eFluor450 | Thermo Fisher Scientific | 48-1619-42 | HP-3G10 | 1,25 µg/ml |
| CD183 (CXCR3) PE/Cy7 | BioLegend | 353720 | G025H7 | 2 µg/ml |
| CD185 (CXCR5) BV750 | BD Bioscience | 747111 | RF8B2 | 1,2 µg/ml |
| CD195 (CCR5) BUV563 | BD Bioscience | 741401 | 2D7/CCR5 | 5 µg/ml |

| | | | | |
|---|---|---|---|---|
| CD196 (CCR6) BV711 | BioLegend | 353436 | G034E3 | 0,6 µg/ml |
| CD197 (CCR7) BV421 | BioLegend | 353208 | G043H7 | 3,5 µg/ml |
| CD279 (PD-1) BV785 | BioLegend | 329930 | EH12.2H7 | 5 µg/ml |
| HLA-DR APC Fire750 | BioLegend | 307658 | L243 | 1,2 µg/ml |
| IgD BV480 | BD Bioscience | 566138 | IA6-2 | 4 µl from a 1:10 pre-dilution in 100 µl |
| IgG BV605 | BD Bioscience | 563246 | G18-145 | 1 µg/ml |
| IgM BV570 | BioLegend | 314517 | MHM-88 | 2,5 µl in 100 µl |
| TCRγδ PerCPeFluor710 | Thermo Fisher Scientific | 46-9959-42 | B1.1 | 4 µg/ml |

| Materials | Company | Catalog number |
|---|---|---|
| Human True StainFcX | BioLegend | 422302 |
| True Stain Monocyte block | BioLegend | 426103 |
| Cell Staining Buffer | BioLegend | 420201 |
| True-Nuclear Transcription Factor Buffer Set | BioLegend | 424401 |
| Human True StainFcX | BioLegend | 422302 |
| DNase | Merck-Millipore | 260913-10MU |
| FCS | Sigma | |
| RPMI1640 | Sigma | R0883 |
| DPBS | Gibco | 14190 |

**Table S4 CombROC mild vs severe**

| Combi-nation | AUC | SE | SP | CutOff | ACC | TN | TP | FN | FP | NPV | PPV | Population |
|---|---|---|---|---|---|---|---|---|---|---|---|---|
| 1 | 0,786 | 0,714 | 1 | 0,532 | 0,882 | 10 | 5 | 2 | 0 | 0,833 | 1 | $CD14^+CD16^+$ monocytes |
| 64 | 0,971 | 1 | 0,9 | 0,34 | 0,941 | 9 | 7 | 0 | 1 | 1 | 0,875 | intermediate effector $CD8^+$ T cells & MAIT cells |
| 263 | 0,971 | 1 | 0,9 | 0,295 | 0,941 | 9 | 7 | 0 | 1 | 1 | 0,875 | $CD8^+$ NKT cells & intermediate effector $CD8^+$ T cells & MAIT cells |
| 308 | 0,971 | 1 | 0,9 | 0,328 | 0,941 | 9 | 7 | 0 | 1 | 1 | 0,875 | $CD8^+HLA\text{-}DR^+CD38^+$ T cells & intermediate effector $CD8^+$ T cells & MAIT cells |
| 344 | 0,986 | 1 | 0,9 | 0,292 | 0,941 | 9 | 7 | 0 | 1 | 1 | 0,875 | cDC1 & intermediate effector $CD8^+$ T cells & MAIT cells |
| 403 | 1 | 1 | 1 | 0,5 | 1 | 10 | 7 | 0 | 0 | 1 | 1 | intermediate effector $CD8^+$ T cells & MAIT cells & NKT cells |

AUC, Area under curve; SE, sensitivity; SP, specificity; ACC, accuracy; TN, true negative; TP, true positives; FN, false negatives; FP, false positives; NPV, negative predictive value; PPV, positive predictive value

Table **S5 CombiROC mild vs moderate**

| Combi-nation | AUC | SE | SP | CutOff | ACC | TN | TP | FN | FP | NPV | PPV | Population |
|---|---|---|---|---|---|---|---|---|---|---|---|---|
| 72 | 0,8 | 0,85 | 0,7 | 0,619 | 0,8 | 7 | 17 | 3 | 3 | 0,7 | 0,85 | MAIT cells & $IgG^+$ $CD95^+$ B cells |
| 429 | 0,885 | 0,65 | 1 | 0,747 | 0,767 | 10 | 13 | 7 | 0 | 0,588 | 1 | MAIT cells & plasmablasts & $IgG^+CD95^+$ B cells |
| 1436 | 0,935 | 0,8 | 1 | 0,764 | 0,867 | 10 | 16 | 4 | 0 | 0,714 | 1 | MAIT cells & plasmablasts & $IgG^+$ $CD95^+$ B cells & NKT cells |
| 2976 | 0,94 | 0,9 | 0,9 | 0,704 | 0,9 | 9 | 18 | 2 | 1 | 0,818 | 0,947 | $CD8^+$ NKT cells & MAIT cells & plasmablasts & $IgG^+$ $CD95^+$ B cells & NKT cells |
| 5282 | 0,975 | 0,9 | 1 | 0,804 | 0,933 | 10 | 18 | 2 | 0 | 0,833 | 1 | $CD4^+$ $T_{emra}$ cells & $CD8^+$ $HLA\text{-}DR^+$ $CD38^+$ T cells & $IgG^+$ $CD95^+$ B cells & neutrophils & plasmablasts & NKT cells |
| 8384 | 0,98 | 0,95 | 1 | 0,749 | 0,967 | 10 | 19 | 1 | 0 | 0,909 | 1 | $CD4^+$ $T_{emra}$ cells & $CD8^+$ NKT cells & $CD8^+HLA\text{-}DR^+CD38^+$ T cells & $IgG^+CD95^+$ B cells & neutrophils & plasmablasts & NKT cells |

### Table S6 CombiROC mild vs moderate & severe

| Combi-nation | AUC | SE | SP | CutOff | ACC | TN | TP | FN | FP | NPV | PPV | Population |
|---|---|---|---|---|---|---|---|---|---|---|---|---|
| 76 | 0,811 | 0,852 | 0,7 | 0,621 | 0,811 | 7 | 23 | 4 | 3 | 0,636 | 0,885 | MAIT cells & plasmablasts |
| 429 | 0,911 | 0,704 | 1 | 0,765 | 0,784 | 10 | 19 | 8 | 0 | 0,556 | 1 | MAIT cells & plasmablasts & $IgG^+CD95^+$ B cells |
| 1436 | 0,93 | 0,815 | 1 | 0,776 | 0,865 | 10 | 22 | 5 | 0 | 0,667 | 1 | MAIT cells & plasmablasts & $IgG^+$ $CD95^+$ B cells & NKT cells |
| 2449 | 0,959 | 0,926 | 0,9 | 0,538 | 0,919 | 9 | 25 | 2 | 1 | 0,818 | 0,962 | $CD4^+$ $T_{emra}$ cells & $CD8^+$ $HLA\text{-}DR^+$ $CD38^+$ T cells & $IgG^+$ $CD95^+$ B cells & neutrophils & plasmablasts |
| 3312 | 0,922 | 0,815 | 1 | 0,796 | 0,865 | 10 | 22 | 5 | 0 | 0,667 | 1 | MAIT cells & plasmablasts & $IgG^+$ $CD95^+$ B cells & NKT cells & cDC1 |
| 5152 | 0,978 | 0,926 | 1 | 0,817 | 0,946 | 10 | 25 | 2 | 0 | 0,833 | 1 | $CD4^+$ $T_{emra}$ cells & $CD8^+$ $HLA\text{-}DR^+CD38^+$ T cells & $IgG^+CD95^+$ B cells & neutrophils & plasmablasts & cDC1 |
| 8762 | 0,981 | 0,926 | 1 | 0,818 | 0,946 | 10 | 25 | 2 | 0 | 0,833 | 1 | $CD4^+$ $T_{emra}$ cells & $CD8^+$ $HLA\text{-}DR^+CD38^+$ T cells & $IgG^+CD95^+$ B cells & neutrophils & plasmablasts & cDC1 & NKT cells |
